# Effect of *APOE* ε4 and its modification by sociodemographic characteristics on cognitive measures in South Asians from LASI-DAD

**DOI:** 10.1101/2024.01.04.24300840

**Authors:** Yi Zhe Wang, Wei Zhao, Priya Moorjani, Alden L. Gross, Xiang Zhou, Aparajit B. Dey, Jinkook Lee, Jennifer A. Smith, Sharon L.R. Kardia

**Affiliations:** Department of Epidemiology, School of Public Health, University of Michigan, 1415 Washington Heights, Ann Arbor, MI 48109, United States of America; Survey Research Center, Institute for Social Research, University of Michigan, 426 Thompson Street, Ann Arbor, MI 48109, United States of America; Department of Molecular and Cell Biology, University of California, Berkeley, 176 Stanley Hall #3220, Berkeley, CA 94720, United States of America; Center for Computational Biology, University of California, Berkeley, 176 Stanley Hall #3220, Berkeley, CA 94720, United States of America; Johns Hopkins Bloomberg School of Public Health, 2024 E Monument St, Baltimore, MD 21205, United States of America; Department of Biostatistics, School of Public Health, University of Michigan, 1415 Washington Heights, Ann Arbor, MI 48109, United States of America; Department of Geriatric Medicine, All India Institute of Medical Sciences, Ansari Nagar, New Delhi 110029, India; Department of Economics and Center for Social Research, University of Southern California, 635 Downey Way, VPD 305, Los Angeles, CA 90089, United States of America

**Keywords:** *APOE*, cognitive function, dementia, sociodemographic characteristics, interaction, South Asian, India

## Abstract

**BACKGROUND:** We investigated effects of *APOE* ε4 and its interactions with sociodemographic characteristics on cognitive measures in 2,563 South Asians from the Diagnostic Assessment of Dementia for the Longitudinal Aging Study of India (LASI-DAD).

**METHODS:** Linear regression models were used to assess the association between *APOE* ε4 and global- and domain-specific cognitive function. Effect modification by age, sex, and education were explored using cross-product interaction terms and subgroup analyses.

**RESULTS:** *APOE* ε4 was inversely associated with most cognitive measures. This association was stronger in the older age group (age >68) for general cognitive function, orientation, and language/fluency, as well as in females for memory and language/fluency. Interaction between *APOE* ε4 and education was mostly nonsignificant.

**DISCUSSION:** *APOE* ε4 is associated with lower cognitive function in South Asians from India, with a more pronounced impact observed in females and older individuals.

## 1 INTRODUCTION

Dementia, a group of neurological disorders characterized by impaired cognitive function, is a leading cause of death, disability, and dependency among older adults worldwide [1,2]. Dementia and cognitive function have significant genetic components, with the ε4 allele of the apolipoprotein E (*APOE*) gene being the strongest genetic risk factor for late-onset Alzheimer’s disease (AD) in many populations [3–5]. For instance, compared to the most common and risk-neutral ε3/ε3 genotype in European ancestry populations, having one ε4 allele is associated with an increase of 2-3 times in the risk of AD, while having two ε4 alleles is associated with an increase in risk of up to 15 times [6]. Conversely, the ε2 allele has been generally associated with reduced AD risk. These *APOE* alleles have also been linked, to a lesser extent, with global cognition and performance in specific cognitive domains including episodic memory, executive function, and verbal fluency in individuals without dementia [7].

Sociodemographic factors such as age, sex, and education are also associated with cognitive function in older adults[8] and contribute to the risk of dementia [9,10]. A growing body of literature has explored the potential interaction between *APOE* ε4 and these sociodemographic characteristics, yet the evidence remains inconclusive. Specifically, some studies report that the effect of *APOE* ε4 on AD may be more pronounced in young-old adults (e.g., 60-70 years) than in those who are older (>70 years) [6,11,12], females than males [6,13–15], and in those with lower education [16,17]. Similar findings are observed for the interactions of *APOE* ε4 with sex and education with cognitive function as the outcome [18–21]. However, in contrast to AD, the effect of *APOE* ε4 on cognitive function appears to increase progressively with age [22,23]. Other studies, however, have failed to detect any interactions between *APOE* ε4 and age, sex, or education in relation to cognitive function and dementia [24–27]. Therefore, it is still unclear to what extent these sociodemographic factors modulate *APOE* ε4 associations.

To date, the vast majority of genomic studies on cognitive function and dementia have been conducted in European ancestry populations [28–32]. India is the most populous country in the world with over 4.1 million people estimated to have dementia [2], yet Indian and South Asian populations in general are rarely represented in genomic studies. This lack of representation is concerning as differences in allele frequencies and linkage disequilibrium (LD) patterns across ancestry groups could result in varying genetic effects and associations with disease [33]. Indeed, there is some evidence that the effect of *APOE* ε4 on cognitive function and dementia differs across ancestries and race/ethnicities [34]. An early meta-analysis of clinical and autopsy-based studies, for example, found that the association between *APOE* ε4 and AD was weaker among African Americans and Hispanics compared to Caucasians, but stronger in Japanese [6]. In India, the few studies that have investigated the relationship between *APOE* ε4 and AD suggest that its effect size is similar to that observed in populations of European ancestry [35–38]. However, prior studies in India have primarily relied on geographically restricted samples. Furthermore, India possesses unique cultural, social, and environmental characteristics, such as dietary practices and pathogen or toxicant exposures, that further distinguish it from populations of European ancestry. These closely intertwined factors may be associated with cognitive function and dementia and/or may modulate the genetic effect of *APOE* ε4 in various ways. Therefore, large-scale epidemiological studies employing nationally representative data are needed to comprehensively explore the effects of *APOE* ε4 on cognition and dementia and identify potential effect modifiers in Indian/South Asian populations.

The Diagnostic Assessment of Dementia for Longitudinal Aging Study in India (LASI-DAD) is a population-based prospective cohort study that has collected nationally representative data on Indians aged 60 and older to better understand the determinants of late-life cognition, cognitive aging, and dementia. In this study, we examined the effect of the *APOE* ε4 allele and its potential modification by sociodemographic factors including age, sex, and education on cognitive measures in LASI-DAD.

## 2 METHODS

### 2.1 Study population

The Longitudinal Aging Study of India (LASI) is a nationally representative sample of more than 73,000 adults from India aged 45 years or older [39]. LASI-DAD is an add-on study of late-life cognition and dementia, in which a subsample of community-residing LASI respondents aged 60 or older were administered in-depth cognitive tests and informant interviews, adopting the Harmonized Cognitive Assessment Protocol (HCAP) developed by the Health and Retirement Study (HRS) to be suitable for the local context and population characteristics [40]. A two-stage stratified random sampling method was used in LASI-DAD to ensure a sample with a broad distribution of cognitive ability including respondents with dementia and mild cognitive impairment (MCI). The respondents were stratified according to their state of residence and their risk of cognitive impairment based on their performance on the cognitive tests and proxy interview (i.e., interview with a family member when the respondent is incapable of participating in the interview) in the main LASI study. Next, within each state, a predetermined sample size proportional to the state’s population size was used to randomly select an equal number of respondents at high and low risk of cognitive impairment [40]. The baseline (Wave 1) LASI-DAD sample consists of 4,096 older adults aged 60 or older, representing >600 ethnically and geographically diverse areas within 18 states and union territories.

### 2.2 Measures

#### 2.2.1 Whole Genome Sequencing

Whole genome sequencing (WGS) was performed on a total of 2,762 LASI-DAD participants who consented to blood sample collection at MedGenome, Inc (Bangalore, India) at an average read depth of 30. Genotype calling and quality control (QC) on the raw LASI-DAD WGS data were performed at the Genome Center for Alzheimer’s Disease (GCAD) at the University of Pennsylvania [41,42]. Briefly, sample-level quality control included checks for low coverage, sample contamination, sex discrepancies, concordance with previous genotype data, and duplicates [41]. After excluding control samples and samples with low quality and/or unresolved identity, a total of 2,680 samples were retained in the analysis. At the genotype level, each genotype was evaluated and set to missing if read depth was less than 10 (DP<10) or genotype quality score was less than 20 (GQ<20). At the variant level, a variant was excluded if it was monomorphic, was above the 99.8% Variant Quality Score Recalibration (VQSR) Tranche (the quality score was beyond the range that contains 99.8% of true variants), had a call rate ≤ 80%, or had an average mean depth > 500 reads. We further removed variants that were in low complexity regions identified with mdust [43]. After quality control and filtering, we retained a total of 71,109,961 autosomal bi-allelic variants that include 66,204,161 single nucleotide polymorphisms (SNPs) and 4,905,800 indels. Genetic principal components (PCs) were calculated using PCAir [44] to take into account the relatedness in the samples. Specifically, the PCs were first calculated in a set of unrelated individuals (kinship cutoff=0.044) and then projected to all other individuals. For this analysis, we included 2,607 individuals that are not related at 3rd degree or above. The top 10 PCs were included in all analyses to adjust for population stratification.

#### 2.2.2 *APOE* Genotyping

*APOE* ε2, ε3, and ε4 alleles are defined by two SNPs (rs429358 and rs7412) that lead to changes in the amino acid sequence at positions 112 and 158. Direct genotyping for these *APOE* SNPs was performed on 2,754 LASI-DAD participants using TaqMan assays (Applied Biosystems, Foster City, CA). The resulting genotypes of these two SNPs were then combined to form six possible *APOE* genotypes: ε2/ε2, ε2/ε3, ε2/ε4, ε3/ε3, ε3/4, and ε4/ε4. In our primary analyses, we dichotomized *APOE* genotypes based on the presence or absence of the ε4 alleles and classified participants as *APOE* ε4 carriers (i.e., ε3/4, and ε4/ε4) or noncarriers (i.e., ε2/ε2, ε2/ε3, and ε3/ε3). We focused on the 2,590 unrelated participants who had both WGS and *APOE* genotyping data. Due to the potential opposing effects of the ε4 and ε2 alleles on cognitive measures, individuals with the *APOE* ε2/ε4 genotype were further excluded from the analyses (N= 27, 1.0%). This left 2,563 participants for inclusion in the primary analyses. A study flow chart is presented in **Supplementary Figure 1**.

#### 2.2.3 Cognitive measures

We examined a total of seven cognitive measures, including the Hindi Mental State Examination (HMSE) score, five broad cognitive domain scores (orientation, memory, executive functioning, language/fluency, and visuospatial), and a general cognitive function summary score constructed from the five domains. The HMSE score is the Hindi version of the Mini-Mental State Examination (MMSE), a widely used cognitive screening instrument [45]. The HMSE score ranges from 0 to 30, with higher scores indicating better cognitive functioning. General cognitive function and domain-specific cognitive performance were derived using factor analysis of the LASI-DAD cognitive battery, as described by Gross et al.[46] These scores were internally standardized to a N(0,1) distribution in the entire sample of 4,096 LASI-DAD participants.

#### 2.2.4 Covariates and effect modifiers

We examined several sociodemographic and socioeconomic characteristics that were self-reported by the participants at baseline as covariates. Sociodemographic factors included age, sex, educational level according to a three-tier simplified version of the 1997 International Standard Classification of Education (ISCED-97) codes (less than secondary education, upper secondary education and vocational training, or tertiary education), urban/rural residence, literacy (can read or write), and state of residence. We also included caste and per capita household consumption (in rupees) as measures of socioeconomic status. Respondents self-reported their caste and were classified into four official categories recognized by the Indian government: scheduled caste, scheduled tribe, other backward class (OBC), or no caste/other caste. Per capita household consumption was used to capture the economic status of the respondents and was calculated by taking total household consumption over the previous year for food, household utilities, fees, durable goods, education, healthcare, discretionary spending, transit, and remittances divided by the number of people in the household.

Age, sex, and education were also assessed as potential effect modifiers of the association between *APOE* ε4 and cognitive function. We examined the interaction between ε4 and education using the three-level categorical education variable described above. Nearly half of our sample (∼48.7%) had never attended school, which aligns with the 56.5% illiteracy rate among older adults in India [47]. Thus, we further investigated the interaction between *APOE* ε4 and a dichotomous education variable, distinguishing between individuals with no formal education and those with any formal education (0 years of education vs. >0 years of education groups). Furthermore, to facilitate interaction and subgroup analyses, age was dichotomized at the sample median (≤ 68 years vs. >68 years) when analyzed as an effect modifier.

### 2.3 Statistical analysis

The distribution of *APOE* genotypes and allele frequencies were calculated for the full study sample, and Hardy-Weinberg equilibrium was assessed. *APOE* genotype and allele frequencies were additionally estimated by 5-year age group ranging from 60 years (smallest age of the sample) to ≥ 85 years. We compared and tested whether the *APOE* ε4 allele frequency differed by age to assess a potential selective survival bias. Descriptive statistics of the analytic sample were compared by *APOE* ε4 carrier status using t-test, Wilcoxon rank-sum test, and Pearson chi-square test as appropriate.

We first examined the association between *APOE* ε4 (carrier vs. non-carrier) and cognitive measures in the full analytic sample using linear regression models. We then tested the interaction between *APOE* ε4 and age, sex, or education by including the two independent variables and their cross-product term into the same model. We further conducted age-, sex-, and education-stratified analyses to estimate the ε4 effects separately within each subgroup, regardless of whether an interaction was detected. We employed two sets of models for all analyses. Model 1 adjusted for age (if applicable), sex (if applicable), state of residence, and the top 10 genetic PCs. Model 2 additionally adjusted for social- and economic-related variables including educational level (if applicable), literacy, urban/rural residence, caste, and per capita household consumption.

As a sensitivity analysis, we repeated all analyses to examine the effects of specific *APOE* genotypes and their modification by age, sex, and education on cognitive measures. In this analysis, we excluded individuals with the ε2/ε4 (protective/risk allele) and ε2/ε2 (low frequency; N=5) genotypes, and focused on the remaining four possible *APOE* genotypes: ε3/ε3 (reference group), ε2/ε3, ε3/ε4, and ε4/ε4. We did not test the interaction between *APOE* genotypes and the three-tier education variable due to the smaller subgroup sample sizes.

## 3 RESULTS

### 3.1 Sample characteristics

**Supplementary Table 1** provides the distribution of *APOE* genotypes and alleles in the full study population, as well as by 5-year age intervals ranging from 60 years to ≥ 85 years. The allele frequencies of *APOE* ε2, ε3, and ε4 were 0.048, 0.844, and 0.108, respectively. There was no significant difference in the frequency of the ε4 allele by age when compared across 5-year age groups (p = 0.813) (**Supplementary Table 1 and Supplementary Figure 2**). About 19% (n = 497) of the participants were classified as *APOE* ε4 carriers (18% ε3/ε4 heterozygotes and 1% ε4/ε4 homozygotes). Genotype frequencies observed in the full study sample were in Hardy-Weinberg equilibrium (p = 0.667).

Descriptive statistics for sociodemographic characteristics and cognitive measures by *APOE* ε4 carrier status are shown in **Table 1**. The mean age of the participants was 69.56 years (SD 7.29) and approximately 53% (n = 1,358) were female. Seventy-five percent (n=1,926) had less than lower secondary education, with 48.7% (n= 1,249) participants having no formal education. Twenty-one percent (n= 542) had upper secondary or vocational training, and 4% (n=95) had tertiary education. *APOE* ε4 carriers and noncarriers did not differ with respect to age, sex, and state of residence. However, compared to *APOE* ε4 noncarriers, ε4 carriers were more likely to have less than upper secondary education, reside in rural areas, and score lower on all cognitive measures except visuospatial functioning. Significant differences in caste and per capita household consumption were also found between *APOE* ε4 carriers and noncarriers.

**Table 1.** Descriptive statistics by *APOE* ε4 carrier status among 2,563 participants in LASI-DAD.

| | Total<br>(N = 2,563) | *APOE* $\epsilon 4$<br>noncarrier<br>(n = 2,066) | *APOE* $\epsilon 4$ carrier<br>(n = 497) | P value |
| --- | --- | --- | --- | --- |
| Age, mean (SD), years | 69.56 (7.29) | 69.55 (7.27) | 69.59 (7.39) | 0.996<sup>a</sup> |
| Female sex | 1358 (53.0%) | 1098 (53.1%) | 260 (52.3%) | 0.777<sup>b</sup> |
| State of residence |  |  |  | 0.084<sup>b</sup> |
| Assam | 68 (2.7%) | 47 (2.3%) | 21 (4.2%) |  |
| Bihar | 129 (5.0%) | 101 (4.9%) | 28 (5.6%) |  |
| Delhi | 119 (4.6%) | 94 (4.5%) | 25 (5.0%) |  |
| Gujarat | 220 (8.6%) | 174 (8.4%) | 46 (9.3%) |  |
| Haryana | 165 (6.4%) | 137 (6.6%) | 28 (5.6%) |  |
| Jammu & Kashmir | 85 (3.3%) | 73 (3.5%) | 12 (2.4%) |  |
| Karnataka | 115 (4.5%) | 98 (4.7%) | 17 (3.4%) |  |
| Kerala | 250 (9.8%) | 208 (10.1%) | 42 (8.5%) |  |
| Madhya Pradesh | 73 (2.8%) | 59 (2.9%) | 14 (2.8%) |  |
| Maharashtra | 173 (6.7%) | 131 (6.3%) | 42 (8.5%) |  |
| Orissa | 206 (8.0%) | 156 (7.6%) | 50 (10.1%) |  |
| Punjab | 105 (4.1%) | 83 (4.0%) | 22 (4.4%) |  |
| Rajasthan | 144 (5.6%) | 124 (6.0%) | 20 (4.0%) |  |
| Tamil Nadu | 155 (6.0%) | 125 (6.1%) | 30 (6.0%) |  |
| Telangana | 164 (6.4%) | 142 (6.9%) | 22 (4.4%) |  |
| Uttar Pradesh | 148 (5.8%) | 114 (5.5%) | 34 (6.8%) |  |
| Uttaranchal | 63 (2.5%) | 51 (2.5%) | 12 (2.4%) |  |
| West Bengal | 181 (7.1%) | 149 (7.2%) | 32 (6.4%) |  |
| Education |  |  |  | 0.024<sup>b</sup> |
| Less than lower secondary | 1926 (75.1%) | 1529 (74.0%) | 397 (79.9%) |  |
| Upper secondary or vocational training | 542 (21.1%) | 458 (22.2%) | 84 (16.9%) |  |
| Tertiary | 95 (3.7%) | 79 (3.8%) | 16 (3.2%) |  |
| Education (dichotomous) |  |  |  | 0.067<sup>b</sup> |
| No formal education (0 years of education) | 1249 (48.7%) | 988 (47.8%) | 261 (52.5%) |  |
| Some level of education (>0 years of education) | 1314 (51.3%) | 1078 (52.2%) | 236 (47.5%) |  |
| Rural residence | 1625 (63.4%) | 1283 (62.1%) | 342 (68.8%) | 0.006<sup>b</sup> |
| Illiteracy (cannot read or write) | 1455 (56.8%) | 1157 (56.0%) | 298 (60.0%) | 0.121<sup>b</sup> |
| Caste (n = 2,550) |  |  |  | 0.001<sup>b</sup> |
| Scheduled caste | 467 (18.2%) | 376 (18.2%) | 91 (18.3%) |  |
| Scheduled tribe | 99 (3.9%) | 66 (3.2%) | 33 (6.6%) |  |
| Other backward class | 1120 (43.7%) | 896 (43.4%) | 224 (45.1%) |  |
| No caste or other caste | 864 (33.7%) | 717 (34.7%) | 147 (29.6%) |  |
| Per capita household consumption in quintiles, rupees (n = 2,561) |  |  |  | 0.013<sup>b</sup> |
| First ($\leq 21,973$) | 512 (20.0%) | 388 (18.8%) | 124 (24.9%) | |
| Second (21,973 – 30,820) | 514 (20.1%) | 416 (20.1%) | 98 (19.7%) |  |
| Third (30,820 – 42,489) | 509 (19.9%) | 406 (19.7%) | 103 (20.7%) |  |
| Fourth (42,489 – 62,668) | 514 (20.1%) | 431 (20.9%) | 83 (16.7%) |  |
| Fifth ($\geq 65,206$) | 512 (20.0%) | 423 (20.5%) | 89 (17.9%) | |
| HMSE score, mean (SD) | 22.69 (5.38) | 22.90 (5.17) | 21.79 (6.08) | 0.002<sup>a</sup> |
| General cognitive function, mean (SD) | 0.005 (0.91) | 0.036 (0.90) | -0.124 (0.98) | 0.001<sup>c</sup> |
| Cognitive domain scores, mean (SD) |  |  |  |  |
| Orientation | -0.025 (0.79) | 0.003 (0.78) | -0.143 (0.85) | 4.84E-04<sup>c</sup> |
| Executive function | -0.006 (0.9) | 0.019 (0.89) | -0.112 (0.92) | 0.004<sup>c</sup> |
| Language/fluency | -0.032 (0.8) | -0.006 (0.78) | -0.138 (0.88) | 0.002<sup>c</sup> |
| Memory | 0.021 (0.93) | 0.053 (0.93) | -0.112 (0.96) | 0.001<sup>c</sup> |
| Visuospatial function | 0.029 (0.83) | 0.039 (0.82) | -0.008 (0.84) | 0.261<sup>c</sup> |

Abbreviations: *APOE* = apolipoprotein E; LASI-DAD = Diagnostic Assessment of Dementia for the Longitudinal Aging Study of India; SD = standard deviation. HMSE = Hindi Mental State Examination.
Note: Individuals with the *APOE* $\epsilon 2/\epsilon 4$ genotype were excluded from the sample. N (%) are reported unless otherwise specified.
<sup>a</sup> p-value calculated from Wilcoxon rank sum test.
<sup>b</sup> p-value calculated from Chi-square test.
<sup>c</sup> p-value calculated from t-test.

**Supplementary Table 2** provides the correlation among the seven cognitive measures examined in this study. The correlation among the five cognitive domain scores ranged from 0.491 to 0.733. The correlations between the five domain scores and general cognitive function and HMSE ranged from 0.733 to 0.948 and from 0.560 to 0.856, respectively.

### 3.2 *APOE* ε4 and sociodemographic characteristic associations with cognitive measures

**Table 2** presents the associations between *APOE* ε4 and cognitive measures in the full sample. All effect estimations were in the expected direction, with ε4 carriers having lower cognitive function. In models adjusting for age, sex, state of residence, and the top 10 genetic PCs (Model 1), ε4 carrier status was associated with all cognitive measures except visuospatial functioning (p < 0.05). Specifically, being an ε4 carrier was associated with a 0.095 – 0.142 SD decrease in cognitive scores and a 0.978 points reduction in HMSE score. The associations were slightly attenuated after further controlling for education, literacy, urban/rural residence, caste, and per capita household consumption in Model 2. Nevertheless, the association between *APOE* ε4 and HMSE score, general cognitive function, orientation, and memory remained significant (p < 0.05).

**Table 2.** Effects of *APOE* ε4 carrier status on cognitive measures in LASI-DAD.

| Cognitive Measure | Model 1<br>(n = 2,563) |  |  | Model 2<br>(n = 2,548) |  |  |
| --- | --- | --- | --- | --- | --- | --- |
| | Beta | P value | $\Delta R^2$ | Beta | P value | $\Delta R^2$ |
| HMSE score | **-0.978** | **2.44E-05** | 0.51% | **-0.710** | **0.001** | 0.27% |
| General cognitive function | **-0.142** | **1.69E-04** | 0.37% | **-0.079** | **0.006** | 0.11% |
| Executive function | **-0.117** | **0.002** | 0.26% | -0.057 | 0.054 | 0.06% |
| Orientation | **-0.141** | **2.86E-05** | 0.49% | **-0.096** | **0.001** | 0.22% |
| Language/fluency | **-0.095** | **0.005** | 0.21% | -0.052 | 0.064 | 0.07% |
| Memory | **-0.134** | **0.001** | 0.32% | **-0.087** | **0.019** | 0.13% |
| Visuospatial function | -0.063 | 0.097 | 0.09% | -0.019 | 0.576 | 0.01% |

Abbreviations: APOE = apolipoprotein E; LASI-DAD = Diagnostic Assessment of Dementia for the Longitudinal Aging Study of India; HMSE = Hindi Mental State Examination.
Model 1 adjusted for age, sex, state of residence, and the top 10 genetic PCs.
Model 2 adjusted for age, sex, state of residence, top 10 genetic PCs, education (upper secondary or vocational training and tertiary education), literacy, urban/rural residence, caste, and quintiles of per capita household consumption.
$\Delta R^2$ represents the change in $R^2$ when *APOE* ε4 carrier status was added to the corresponding regression models.
Beta coefficient and p-value in bold indicates statistically significant association at $p < 0.05$ .

Effect estimates and p-values for all other covariates in Models 1 and 2 can be found in **Supplementary Tables 3 and 4**, respectively. In Model 1, older age was inversely associated with all cognitive measures, while male sex was positively associated with all cognitive measures. These associations stayed largely consistent in Model 2. Education was also independently associated with all cognitive measures. Compared to individuals with less than lower secondary education, those with upper secondary or vocational training education showed a 0.179-0.458 SD increase in cognitive scores and a 1.480 point increase in HMSE score, while those with tertiary education showed a 0.327-0.893 SD increase in cognitive scores and a 2.392 points increase in HMSE score.

Collectively, *APOE* ε4 carrier status and other covariates in Model 1 explained approximately 18.6% (visuospatial function) to 33.9% (general cognitive function) of the total variance in cognitive measures. Specifically, ε4 carrier status alone contributed between 0.09% (visuospatial function) and 0.51% (HMSE score) of the explained variance. The total variance explained increased to 37.9-62.4% when education, literacy, urban/rural residence, caste, and per capita household consumption were added to the model (Model 2). In this further adjusted model, the individual contribution of ε4 carrier status to the total variance explained ranged from 0.01% to 0.27%.

### 3.3 Effect modification of age, sex, and education on the associations between *APOE* **ε**4 and cognitive measures

We next examined whether the associations of *APOE* ε4 with cognitive measures were modified by age, sex, and education. **Table 3** provides the results from testing the *APOE* ε4 × age > 68 years interaction term. We found a significant *APOE* ε4 by age interaction on general cognitive function, orientation, and language/fluency in Model 2, with the associations being more pronounced in the > 68 years age group (**Figure 1**). Results from age-stratified analysis suggest that the detected interactions were mainly driven by the significant ε4 effects on cognitive function, orientation, and language/fluency in the > 68 years age group that were not observed in the ≤ 68 years age group (**Table 4**). We also observed similar results for several of the other cognitive measures, where the ε4 effect was significant solely in the older age group.

**Figure 1.**
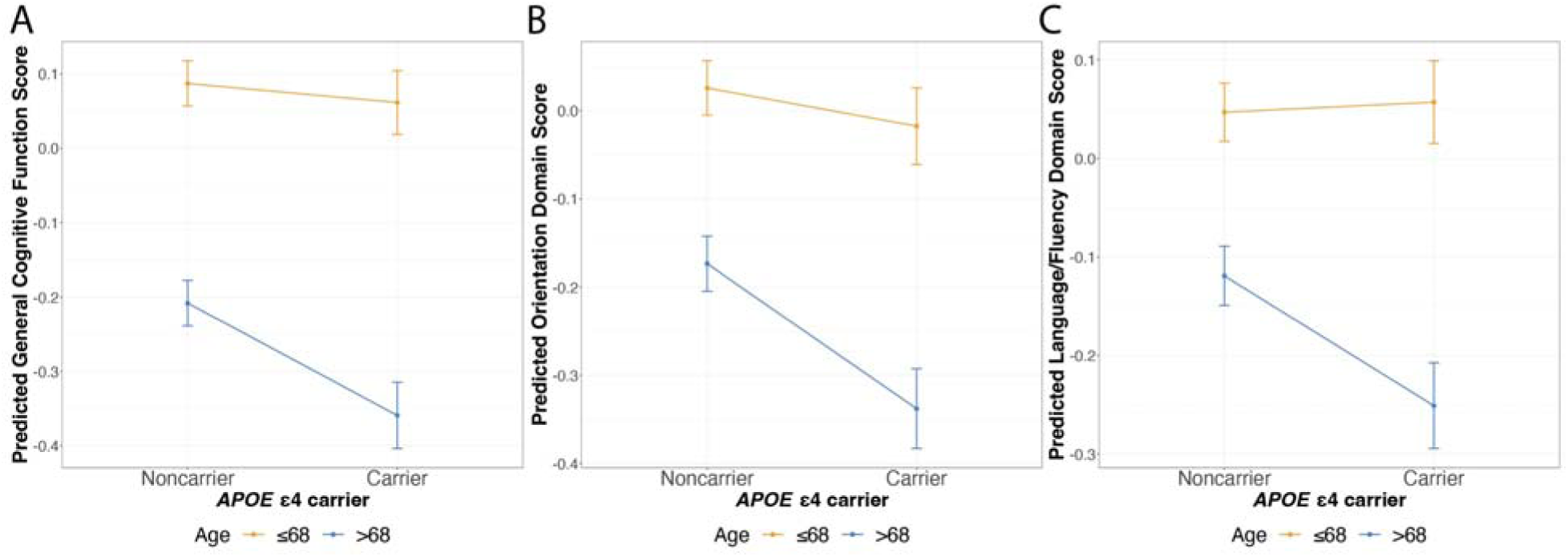
Plots of significant two-way interactions between *APOE* ε4 and age on cognitive measures. Predicted general cognitive function score (A), orientation score (B), and language/fluency score (C) of *APOE* ε4 carriers and noncarriers are shown with standard error bars by age group. Cognitive measures are predicted using models adjusted for age (age >68), sex (male), state of residence, top 10 genetic PCs, education (upper secondary or vocational training and tertiary education), urban/rural residence, literacy, caste, quintiles of per capita household consumption, and *APOE* ε4 × age >68 (Model 2). Color coding is according to age group. Abbreviations: APOE = apolipoprotein E; PC = principal components.

**Table 3.** Two-way interaction between *APOE* ε4 carrier status and age on cognitive measures.

|  | **Model 1**<br>**(n = 2,563)** |  | **Model 2**<br>**(n = 2,548)** |  |
| --- | --- | --- | --- | --- |
|  | **Beta** | **P value** | **Beta** | **P value** |
| **HMSE score** |  |  |  |  |
| *APOE* ε4 | **-0.825** | **0.010** | -0.425 | 0.140 |
| Age > 68 | **-2.059** | **1.02E-22** | **-1.646** | **5.81E-18** |
| *APOE* ε4 × Age > 68 | -0.416 | 0.375 | -0.678 | 0.110 |
| **General cognitive function\*** |  |  |  |  |
| *APOE* ε4 | **-0.122** | **0.019** | -0.026 | 0.516 |
| Age > 68 | **-0.403** | **1.12E-31** | **-0.295** | **3.12E-29** |
| *APOE* ε4 × Age > 68 | -0.058 | 0.449 | **-0.126** | **0.031** |
| **Executive function** |  |  |  |  |
| *APOE* ε4 | **-0.109** | **0.036** | -0.014 | 0.741 |
| Age > 68 | **-0.352** | **4.49E-25** | **-0.248** | **6.62E-20** |
| *APOE* ε4 × Age > 68 | -0.032 | 0.671 | -0.104 | 0.086 |
| **Orientation\*** |  |  |  |  |
| *APOE* ε4 | **-0.107** | **0.021** | -0.043 | 0.281 |
| Age > 68 | **-0.264** | **2.81E-18** | **-0.199** | **5.52E-14** |
| *APOE* ε4 × Age > 68 | -0.086 | 0.208 | **-0.121** | **0.040** |
| **Language/fluency\*** |  |  |  |  |
| *APOE* ε4 | -0.050 | 0.284 | 0.010 | 0.792 |
| Age > 68 | **-0.237** | **6.66E-15** | **-0.166** | **7.05E-11** |
| *APOE* ε4 × Age > 68 | -0.107 | 0.117 | **-0.142** | **0.012** |
| **Memory** |  |  |  |  |
| *APOE* ε4 | **-0.117** | **0.039** | -0.042 | 0.407 |
| Age > 68 | **-0.441** | **3.40E-32** | **-0.358** | **2.87E-26** |
| *APOE* ε4 × Age > 68 | -0.054 | 0.515 | -0.108 | 0.148 |
| **Visuospatial function** |  |  |  |  |
| *APOE* ε4 | -0.074 | 0.154 | -0.003 | 0.951 |
| Age > 68 | **-0.301** | **6.74E-19** | **-0.216** | **5.09E-13** |
| *APOE* ε4 × Age > 68 | 0.014 | 0.858 | -0.041 | 0.543 |

Abbreviations: *APOE* = apolipoprotein E; LASI-DAD = Diagnostic Assessment of Dementia for the Longitudinal Aging Study of India; HMSE = Hindi Mental State Examination.
Model 1 adjusted for age (age > 68), sex (male), state of residence, top 10 genetic PCs, and *APOE* ε4 × Age > 68.
Model 2 adjusted for age (age > 68), sex (male), state of residence, top 10 genetic PCs, education (upper secondary or vocational training and tertiary education), literacy, urban/rural residence, caste, quintiles of per capita household consumption, and *APOE* ε4 × Age > 68.
Beta coefficient and p-value in bold indicates statistically significant association at $p < 0.05$ .
Asterisk (\*) denotes cognitive measures for which a statistically significant interaction term was observed.

**Table 4.** Associations between *APOE* ε4 carrier status and cognitive measures stratified by the median age of the sample.

| Cognitive Measure | Model 1 |  |  |  | Model 2 |  |  |  |
| --- | --- | --- | --- | --- | --- | --- | --- | --- |
| | Age ≤ 68<br>(n = 1,363) | | Age > 68<br>(n = 1,200) | | Age ≤ 68<br>(n = 1,360) | | Age > 68<br>(n = 1,188) | |
|  | Beta | P value | Beta | P value | Beta | P value | Beta | P value |
| HMSE score | **-0.816** | **0.006** | -<br>1.339 | **3.70E-04** | -0.418 | 0.107 | **-1.194** | **0.001** |
| General cognitive function* | **-0.116** | **0.027** | -<br>0.194 | **0.001** | -0.018 | 0.618 | **-0.167** | **4.28E-04** |
| Executive function | **-0.103** | **0.049** | -<br>0.155 | **0.006** | -0.006 | 0.884 | **-0.131** | **0.006** |
| Orientation* | **-0.103** | **0.020** | -<br>0.206 | **1.00E-04** | -0.043 | 0.244 | **-0.182** | **1.54E-04** |
| Language/fluency* | -0.048 | 0.275 | -<br>0.164 | **0.002** | 0.013 | 0.706 | **-0.140** | **0.003** |
| Memory | -0.103 | 0.072 | -<br>0.186 | **0.002** | -0.027 | 0.588 | **-0.172** | **0.003** |
| Visuospatial function | -0.073 | 0.168 | -<br>0.057 | 0.301 | -0.008 | 0.860 | -0.040 | 0.423 |

Abbreviations: *APOE* = apolipoprotein E; HMSE = Hindi Mental State Examination.
Model 1 adjusted for sex, state of residence, and top 10 genetic PCs.
Model 2 adjusted for sex, state of residence, top 10 genetic PCs, education (upper secondary or vocational training and tertiary education), literacy, urban/rural residence, caste, and quintiles of per capita household consumption.
Beta coefficient and p-value in bold indicates statistically significant association at $p < 0.05$ .
Asterisk (\*) denotes cognitive measures for which a statistically significant interaction term was observed.

**Table 5** presents the results from testing the *APOE* ε4 by sex interaction term. In Model 1, an *APOE* ε4 × male interaction was observed for language/fluency and memory, with the effect of ε4 stronger in females. While the interaction terms were not statistically significant for the other cognitive measures, sex-stratified analysis showed that *APOE* ε4 was associated with all cognitive measures among females except visuospatial functioning, whereas there was no association between ε4 and any cognitive measures among males (**Table 6**). Sex differences in the ε4 effect attenuated in Model 2, but the *APOE* ε4 × male sex interaction term remained statistically significant for both language/fluency and memory (**Figure 2**, **Tables 5 and 6**).

**Figure 2.**
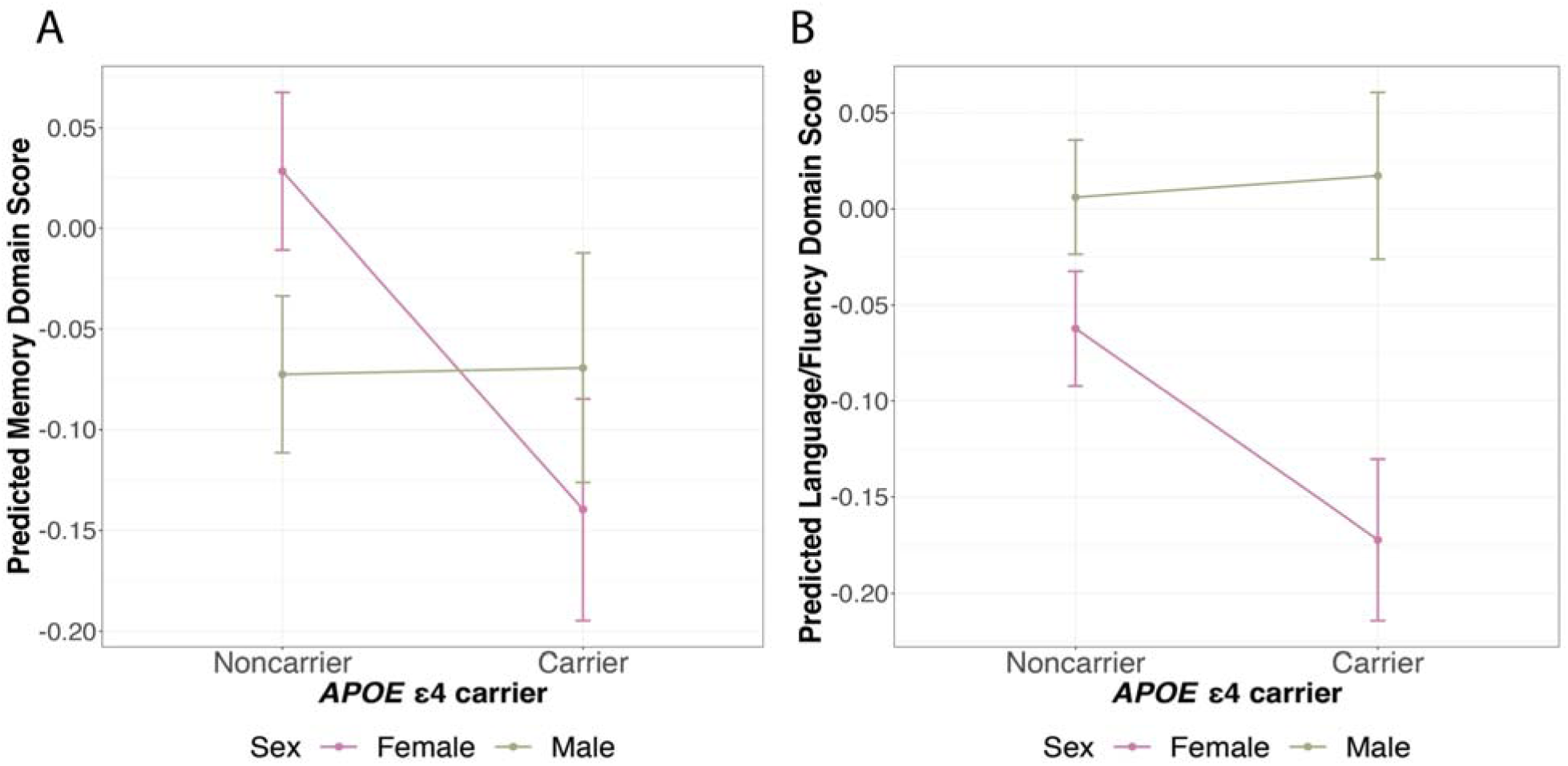
Plots of significant two-way interactions between *APOE* ε4 and sex on cognitive measures. Predicted memory score (A) and language/fluency score (B) of *APOE* ε4 carriers and noncarriers are shown with standard error bars by sex. Cognitive measures are predicted using models adjusted for age, sex (male), state of residence, top 10 genetic PCs, education, urban/rural residence, literacy, caste, quintiles of per capita household consumption, and *APOE* ε4 × male (Model 2). Color coding is according to sex. Abbreviations: APOE = apolipoprotein E; PC = principal components.

**Table 5.** Two-way interaction between *APOE* ε4 carrier status and sex on cognitive measures.

|  | Model 1<br>(n = 2,563) |  | Model 2<br>(n = 2,548) |  |
| --- | --- | --- | --- | --- |
|  | Beta | P value | Beta | P value |
| **HMSE score** |  |  |  |  |
| *APOE* ε4 | **-1.334** | **3.00E-05** | **-0.997** | **0.001** |
| Male | **3.196** | **9.87E-53** | **1.696** | **1.94E-17** |
| *APOE* ε4 × Male | 0.748 | 0.105 | 0.603 | 0.149 |
| **General cognitive function** |  |  |  |  |
| *APOE* ε4 | **-0.199** | **1.34E-04** | **-0.121** | **0.002** |
| Male | **0.611** | **1.53E-70** | **0.232** | **2.20E-17** |
| *APOE* ε4 × Male | 0.120 | 0.112 | 0.089 | 0.121 |
| **Executive function** |  |  |  |  |
| *APOE* ε4 | **-0.128** | **0.014** | -0.056 | 0.177 |
| Male | **0.637** | **6.23E-77** | **0.287** | **7.69E-24** |
| *APOE* ε4 × Male | 0.023 | 0.762 | -0.004 | 0.951 |
| **Orientation** |  |  |  |  |
| *APOE* ε4 | **-0.203** | **1.26E-05** | **-0.145** | **3.50E-04** |
| Male | **0.628** | **2.72E-91** | **0.378** | **6.79E-41** |
| *APOE* ε4 × Male | 0.130 | 0.053 | 0.103 | 0.079 |
| **Language/fluency\*** |  |  |  |  |
| *APOE* ε4 | **-0.166** | **3.88E-04** | **-0.110** | **0.005** |
| Male | **0.355** | **8.35E-32** | **0.068** | **0.010** |
| *APOE* ε4 × Male | **0.149** | **0.027** | **0.121** | **0.031** |
| **Memory\*** |  |  |  |  |
| *APOE* ε4 | **-0.223** | **8.44E-05** | **-0.168** | **0.001** |
| Male | **0.172** | **2.09E-06** | **-0.101** | **0.004** |
| *APOE* ε4 × Male | **0.185** | **0.024** | **0.171** | **0.020** |
| **Visuospatial function** |  |  |  |  |
| *APOE* ε4 | -0.088 | 0.091 | -0.033 | 0.471 |
| Male | **0.441** | **1.92E-38** | **0.148** | **2.66E-06** |
| *APOE* ε4 × Male | 0.054 | 0.478 | 0.030 | 0.647 |

Abbreviations: *APOE* = apolipoprotein E; LASI-DAD = Diagnostic Assessment of Dementia for the Longitudinal Aging Study of India; HMSE = Hindi Mental State Examination.
Model 1 adjusted for age, sex (male), state of residence, top 10 genetic PCs, and *APOE* ε4 × male.
Model 2 adjusted for age, sex (male), state of residence, top 10 genetic PCs, education (upper secondary or vocational training and tertiary education), literacy, urban/rural residence, caste, quintiles of per capita household consumption, and *APOE* ε4 × male.
Beta coefficient and p-value in bold indicates statistically significant association at $p < 0.05$ .
Asterisk (\*) denotes cognitive measures for which a statistically significant interaction term was observed.

**Table 6.** Associations between *APOE* ε4 carrier status and cognitive measures stratified by sex.

| Cognitive Measure | Model 1 |  |  |  | Model 2 |  |  |  |
| --- | --- | --- | --- | --- | --- | --- | --- | --- |
|  | Male<br>(n = 1,205) |  | Female<br>(n = 1,358) |  | Male<br>(n = 1,200) |  | Female<br>(n = 1,348) |  |
|  | Beta | P value | Beta | P value | Beta | P value | Beta | P value |
| HMSE score | -0.522 | 0.105 | -<br>**1.249** | **1.72E-04** | -0.395 | 0.171 | **-0.904** | **0.003** |
| General cognitive function | -0.072 | 0.202 | -<br>**0.187** | **2.09E-04** | -0.037 | 0.386 | **-0.113** | **0.004** |
| Executive function | -0.098 | 0.091 | -<br>**0.118** | **0.016** | -0.060 | 0.186 | -0.052 | 0.191 |
| Orientation | -0.056 | 0.235 | -<br>**0.203** | **2.16E-05** | -0.039 | 0.340 | **-0.144** | **0.001** |
| Language/fluency* | -0.013 | 0.795 | -<br>**0.147** | **0.001** | 0.006 | 0.884 | **-0.092** | **0.017** |
| Memory* | -0.037 | 0.532 | -<br>**0.206** | **2.66E-04** | -0.012 | 0.822 | **-0.150** | **0.003** |
| Visuospatial function | -0.041 | 0.471 | -<br>0.084 | 0.095 | -0.009 | 0.854 | -0.031 | 0.493 |

Abbreviations: APOE = apolipoprotein E; HMSE = Hindi Mental State Examination.
Model 1 adjusted for age, state of residence, and top 10 genetic PCs.
Model 2 adjusted for age, state of residence, top 10 genetic PCs, education (upper secondary or vocational training and tertiary education), literacy, urban/rural residence, caste, and quintiles of per capita household consumption.
Beta coefficient and p-value in bold indicates statistically significant association at $p < 0.05$ .
Asterisk (\*) denotes cognitive measures for which a statistically significant interaction term was observed.

**Supplementary Table 5** provides the results from testing the *APOE* ε4 by education interaction terms. Using the three-level education variable, all interaction terms between *APOE* ε4 and education were not statistically significant except the interaction between ε4 and upper secondary or vocational training on visuospatial function. A closer examination of the education-stratified analysis showed that most significant ε4 associations were predominantly observed in the less than lower secondary education group (**Supplementary Table 6**). Given that this group constituted ∼75% of our sample, with more than 64% of them having no formal education, we further explored the *APOE* ε4 by education interaction by separating our sample into two categories: those with no formal education (0 years of education) and those with any type of formal education (>0 years of education). We found no interaction between *APOE* ε4 and the dichotomous education variable on any of the cognitive measures (**Supplementary Table 7**). However, when we examined the ε4 associations with cognitive measures separately within these two subgroups, we observed that ε4 was associated with all cognitive measures except visuospatial function among individuals without formal education in Model 1 (**Supplementary Table 8**). In contrast, among those who had received some education, ε4 was only associated with HMSE score and orientation domain score. This pattern of association largely persisted in Model 2, except for the loss of significance in the associations between ε4 and executive function and language/fluency among those with no formal education.

### 3.4 Sensitivity analysis

Sensitivity analyses examining *APOE* genotypes showed that the main effect of ε3/ε4 compared to ε3/ε3, as well as its potential modification by age, sex, and education closely mirrored what we observed for ε4 carriers in the primary analyses (**Supplementary Tables 9-15**). Furthermore, even though not statistically significant, we found that the effect of ε4/ε4 on cognitive function was in the expected direction, with lower scores observed among ε4/ε4 compared to ε3/ε3 in the full sample and within age, sex, and education subgroups. Contrary to our expectation based on previous literature, we did not consistently observe a clear dose-response relationship between the number of ε4 alleles and worse cognitive function, as indicated by the smaller magnitude of effect in ε4/ε4 homozygotes compared to ε3/ε4 heterozygotes in the full sample, as well as within subgroups of age ≤68, male sex, and 0 years of education (**Supplementary Tables 9, 11, 13, and 15**). However, we caution that our sample included only a small number of individuals carrying the ε4/ε4 genotype (n=36), which likely contributed to our inability to establish statistically significant associations or interactions.

Finally, results from the sensitivity analyses also revealed an association between *APOE* ε2/ε3 genotype and lower visuospatial function when compared to ε3/ε3 (**Supplementary Table 9**). We did not detect any significant interactions between the ε2/ε3 genotype and age, sex, or education (**Supplementary Table 10, 12, and 14**). Nevertheless, an adverse effect of the ε2/ε3 genotype on visuospatial function was found among females and individuals aged 68 years and older, but not among males and those younger than 68 years (**Supplementary Tables 11 and 13**).

## 4 DISCUSSION

In this large, nationally representative sample of Indians aged 60 and older, *APOE* ε4 carrier status was associated with lower cognitive function, both globally and in multiple specific domains. We also found that females may be more susceptible to ε4 effects on memory and language/fluency, while those aged 68 years and older may be more susceptible to ε4 effects on general cognitive function, orientation, and language/fluency. The ε4 frequency was 10.8% in our sample, which is consistent with the 10.0% ε4 frequency reported in the Genome Aggregation Database (gnomAD) for South Asians [48] and reaffirms the relatively lower ε4 prevalence among South Asians compared to the global frequency of 14% [3]. However, this allele is important as it remains the strongest known genetic risk factor for AD and cognitive function/decline worldwide. Studying its effect on cognitive function contributes to our overall understanding of the genetic architecture of cognitive aging in the Indian population.

We found modest adverse effects of *APOE* ε4 on a broad range of cognitive domains. Direct comparison of ε4 effects on cognition is challenging as methodological factors, including sample size, study population (e.g., participant age range), and study design (e.g., measurement and modeling of cognitive measures), may vary substantially across studies. Nonetheless, the observed domain specificity of ε4 effects on cognition in our study is in line with two previous meta-analyses in cognitively healthy individuals of European ancestry which indicate that ε4 carriers perform worse on measures of global cognition and episodic memory, but not on language/fluency (verbal ability) and visuospatial function [22,49]. However, contrary to these meta-analyses, we did not find an association between ε4 carrier status and executive function after adjusting for sociodemographic and socioeconomic factors. These results also differ slightly from earlier LASI-DAD studies [50,51], which examined genotype chip data on 932 participants and found no significant ε4 effects on cognition, potentially due to inadequate power and a more restricted geographic sample than the current study.

We also found a relatively consistent, though not always statistically significant, pattern across cognitive measures whereby the magnitudes of the ε4 effects on cognition appeared stronger among those at least 68 years than younger individuals. This is consistent with several studies indicating that the negative effect of *APOE* ε4 on cognitive function increases with advancing age [22,52,53]. The molecular mechanisms underlying the potential age-dependent ε4 effects on cognitive function remain unknown. One possibility is the resource modulation hypothesis [53], which posits that effects of common genetic variants, such as *APOE*, may become stronger in older age when neurochemical and anatomical brain resources are declining [54,55]. However, the observed interaction between ε4 and age could also be explained by the increasing presence of preclinical, mildly cognitively impaired, or diagnosed dementia cases with advancing age, as ε4 may be more robustly associated with these cases, creating a seemingly stronger association between ε4 and cognition in older individuals. Not all studies have observed this interaction [49,56]. In fact, one study showed that ε4 effects on global cognitive functioning and episodic memory were inversely related to age, although these correlations were not statistically significant [49]. These inconsistent findings may be partly due to methodologic differences, such as sample size, age range of the participants, and measurements of cognitive function across studies.

We found that the association between *APOE* ε4 and cognition was stronger in females than males for language/fluency and memory, which is consistent with a growing body of literature. For instance, early studies found that the ε4 allele confers a greater risk for AD in women than men [57,58]. These findings were replicated in a seminal meta-analysis that reported that female ε4 carriers had a higher risk of developing AD compared to both female ε4 noncarriers and male ε4 carriers [6]. More recently, studies have shown that ε4 effects on AD biomarkers, including cerebrospinal fluid biomarkers (Amyloid-β and tau), brain metabolism, and brain structure [15,59–61], as well as on cognitive function and decline [18,62,63], are also stronger in females. Although the precise biological mechanisms underlying these observed sex differences remain largely unclear, there is compelling biological plausibility. For example, *APOE* may interact with estrogen through various pathways such that the presence of the ε4 allele exacerbates the female-specific effects of fluctuations/loss of estrogens during perimenopause and postmenopause on AD [64]. Alternatively, the observed sex differences in our sample may, in part, stem from a more pronounced selection bias among males. *APOE* ε4 has pleiotropic effects on cardiovascular disease and mortality, and males may have a higher risk of cardiovascular disease and mortality through *APOE*-related mechanisms and other pathways [19]. Our LASI-DAD sample is generally older and thus more likely to include healthier male ε4 carriers who demonstrate resilience against the detrimental ε4 effects on cognitive function and age-related diseases. Consequently, our study may underestimate the association between ε4 and cognition in males. Indeed, while our findings align with the majority of studies examining sex differences, a few studies have reported conflicting findings [26,65]. For instance, a recent study in UK Biobank participants with white British ancestry found no significant interactions between ε4 and sex on cognitive abilities [65]. Further investigation is required to elucidate the observed sex-specific ε4 effects on cognitive function and dementia in diverse populations.

In our study, as expected, having at least some formal education was associated with better cognitive performance. However, this association did not differ statistically across education groups. Prior research evaluating this interaction in European ancestry samples has yielded conflicting results, with some studies demonstrating that higher education may counteract the negative effects of ε4 on cognitive function and dementia [16,66]. Higher education may provide greater cognitive reserve, allowing individuals to better tolerate age- and dementia-related brain pathologies and maintain cognitive function, thereby mitigating genetic risk and postponing the clinical manifestation of dementia. While we found no evidence of interaction when including a cross-product interaction term between *APOE* ε4 and >0 years of education, education-stratified analysis showed that the magnitude of the association between ε4 and most cognitive measures trended greater in those who never attended school compared to those who did. Lack of significant interactions may have been due, in part, to education being analyzed using a dichotomous variable based on having ever attended school, as some prior studies have suggested that the interaction between ε4 and education may be at the tertiary level [66]. Although we found no interaction between ε4 and tertiary education, it is likely that we lacked statistical power to detect such an interaction due to the limited number of participants with tertiary education (n=95). Further, overall sample size may have limited our power to detect smaller effects. In addition, the complex social hierarchy and high levels of social stratification in India, which we are unable to completely capture, might also influence this interaction.

Sensitivity analyses support our main findings while additionally identifying an association between the ε2/ε3 genotype and lower visuospatial function compared to ε3/ε3. Despite mounting evidence suggesting a protective role of ε2 against AD, previous studies examining its effect on cognition have yielded inconsistent findings. For instance, one study found that ε2 carriers demonstrated worse performance in visuospatial attention in mid-adulthood [67], whereas another study in older Koreans without dementia showed that ε2 carriers performed better on visuospatial measures [68]. Additional research is needed to replicate our findings and elucidate the ε2 effect on cognitive function.

This study has some limitations. First, we were unable to reliably estimate the dosage effect of *APOE* ε4 due to the rarity of ε4 homozygotes (n = 36). Second, LASI-DAD only included participants aged 60 or older, which may have led to selection bias. Our results might underestimate the true association between ε4 and cognition since older adults carrying ε4 and with poorer cognitive function might be less likely to participate due to morbidity or death.

However, ε4 allele frequency did not appear to decrease with age within our sample. This contrasts with findings from prior studies showing a decrease in ε4 frequency with increasing age [69,70]. Notably, our study identified a similar decreasing trend in ε4 up to age 85, at which point a subsequent increase was observed, which may be the result of underlying population structure differences or the small sample size of the oldest age groups. Third, despite adjusting for many potential confounders, there may be residual confounding from lifestyle factors and environmental toxicants. Fourth, all socioeconomic characteristics were self-reported and may be subject to response bias. Fifth, our study was cross-sectional with cognitive function assessed at a single time point. Consequently, we were unable to examine longitudinal ε4 effects. Future studies using longitudinal cognitive measures in LASI-DAD, which are currently being collected, could provide further insights into the intricate interplay among ε4, sociodemographics, and cognitive aging in South Asians.

In conclusion, this is the first study to investigate the effect of *APOE* ε4 and its potential modification by sociodemographic factors on cognitive function in a nationally representative sample of older Indians. Our study provides important initial insights into the relationship between the strongest known genetic risk factor for AD and cognitive function in the South Asians population. Future research using longitudinal cohorts is warranted to provide a better understanding of the genetic influences on cognition and dementia within this genetically and socially diverse population.

## Data Availability

Whole genome sequencing data for the Diagnostic Assessment of Dementia for the Longitudinal Aging Study of India is available from the National Institute on Aging Genetics of Alzheimer's Disease Data Storage Site (NIAGADS), accession number: NG00067. Phenotype data is available at the Gateway to Global Aging website, https://g2aging.org/.

https://dss.niagads.org/datasets/ng00067/

## CONSENT STATEMENT

Informed consent was obtained from all participants included in the study.

## REFERENCES

[1] GBD 2019 Collaborators. Global mortality from dementia: Application of a new method and results from the Global Burden of Disease Study 2019. Alzheimers Dement (N Y) 2021;7:e12200. 10.1002/trc2.12200.

[2] Prince MJ, Wimo A, Guerchet MM, Ali GC, Wu Y-T, Prina M. World Alzheimer Report 2015 - The Global Impact of Dementia: An analysis of prevalence, incidence, cost and trends. London: Alzheimer’s Disease International; 2015.

[3] Liu C-C, Kanekiyo T, Xu H, Bu G. Apolipoprotein E and Alzheimer disease: risk, mechanisms and therapy. Nat Rev Neurol 2013;9:106–18. 10.1038/nrneurol.2012.263.

[4] Reitz C, Mayeux R. Genetics of Alzheimer’s Disease in Caribbean Hispanic and African American Populations. Biological Psychiatry 2014;75:534–41. 10.1016/j.biopsych.2013.06.003.

[5] Miyashita A, Kikuchi M, Hara N, Ikeuchi T. Genetics of Alzheimer’s disease: an East Asian perspective. J Hum Genet 2023;68:115–24. 10.1038/s10038-022-01050-z.

[6] Farrer LA, Cupples LA, Haines JL, Hyman B, Kukull WA, Mayeux R, et al. Effects of age, sex, and ethnicity on the association between apolipoprotein E genotype and Alzheimer disease: a meta-analysis. Jama 1997;278:1349–56.

[7] O’Donoghue MC, Murphy SE, Zamboni G, Nobre AC, Mackay CE. APOE genotype and cognition in healthy individuals at risk of Alzheimer’s disease: A review. Cortex 2018;104:103–23. 10.1016/j.cortex.2018.03.025.

[8] Van Hooren SAH, Valentijn AM, Bosma H, Ponds R, Van Boxtel MPJ, Jolles J. Cognitive functioning in healthy older adults aged 64–81: A cohort study into the effects of age, sex, and education. Aging, Neuropsychology, and Cognition 2007;14:40–54.

[9] Gao S, Hendrie HC, Hall KS, Hui S. The relationships between age, sex, and the incidence of dementia and Alzheimer disease: a meta-analysis. Arch Gen Psychiatry 1998;55:809–15. 10.1001/archpsyc.55.9.809.

[10] Letenneur L, Gilleron V, Commenges D, Helmer C, Orgogozo JM, Dartigues JF. Are sex and educational level independent predictors of dementia and Alzheimer’s disease? Incidence data from the PAQUID project. Journal of Neurology, Neurosurgery & Psychiatry 1999;66:177–83. 10.1136/jnnp.66.2.177.

[11] Saddiki H, Fayosse A, Cognat E, Sabia S, Engelborghs S, Wallon D, et al. Age and the association between apolipoprotein E genotype and Alzheimer disease: A cerebrospinal fluid biomarker–based case–control study. PLoS Med 2020;17:e1003289. 10.1371/journal.pmed.1003289.

[12] Bellou E, Baker E, Leonenko G, Bracher-Smith M, Daunt P, Menzies G, et al. Age-dependent effect of APOE and polygenic component on Alzheimer’s disease. Neurobiology of Aging 2020;93:69–77. 10.1016/j.neurobiolaging.2020.04.024.

[13] Payami H, Montee KR, Kaye JA, Bird TD, Yu CE, Wijsman EM, et al. Alzheimer’s disease, apolipoprotein E4, and gender. JAMA 1994;271:1316–7.

[14] Bretsky PM, Buckwalter JG, Seeman TE, Miller CA, Poirier J, Schellenberg GD, et al. Evidence for an interaction between apolipoprotein E genotype, gender, and Alzheimer disease. Alzheimer Dis Assoc Disord 1999;13:216–21. 10.1097/00002093-199910000-00007.

[15] Altmann A, Tian L, Henderson VW, Greicius MD, Alzheimer’s Disease Neuroimaging Initiative Investigators. Sex modifies the APOE-related risk of developing Alzheimer disease. Ann Neurol 2014;75:563–73. 10.1002/ana.24135.

[16] Wang H-X, Gustafson DR, Kivipelto M, Pedersen NL, Skoog I, Windblad B, et al. Education halves the risk of dementia due to apolipoprotein ε4 allele: a collaborative study from the Swedish Brain Power initiative. Neurobiology of Aging 2012;33:1007.e1–1007.e7. 10.1016/j.neurobiolaging.2011.10.003.

[17] Ma H, Zhou T, Li X, Maraganore D, Heianza Y, Qi L. Early-life educational attainment, APOE ε4 alleles, and incident dementia risk in late life. Geroscience 2022;44:1479–88. 10.1007/s11357-022-00545-z.

[18] Mortensen EL, Høgh P. A gender difference in the association between APOE genotype and age-related cognitive decline. Neurology 2001;57:89–95.

[19] Beydoun MA, Boueiz A, Abougergi MS, Kitner-Triolo MH, Beydoun HA, Resnick SM, et al. Sex differences in the association of the apolipoprotein E epsilon 4 allele with incidence of dementia, cognitive impairment, and decline. Neurobiol Aging 2012;33:720–731.e4. 10.1016/j.neurobiolaging.2010.05.017.

[20] Shadlen M-F, Larson EB, Wang L, Phelan EA, McCormick WC, Jolley L, et al. Education modifies the effect of apolipoprotein epsilon 4 on cognitive decline. Neurobiology of Aging 2005;26:17–24. 10.1016/j.neurobiolaging.2004.03.005.

[21] Seeman TE, Huang M-H, Bretsky P, Crimmins E, Launer L, Guralnik JM. Education and APOE-e4 in Longitudinal Cognitive Decline: MacArthur Studies of Successful Aging. The Journals of Gerontology: Series B 2005;60:P74–83. 10.1093/geronb/60.2.P74.

[22] Wisdom NM, Callahan JL, Hawkins KA. The effects of apolipoprotein E on non-impaired cognitive functioning: a meta-analysis. Neurobiol Aging 2011;32:63–74. 10.1016/j.neurobiolaging.2009.02.003.

[23] Lyall DM, Ward J, Ritchie SJ, Davies G, Cullen B, Celis C, et al. Alzheimer disease genetic risk factor *APOE* e4 and cognitive abilities in 111,739 UK Biobank participants. Age Ageing 2016;45:511–7. 10.1093/ageing/afw068.

[24] Small BJ, Graves AB, McEvoy CL, Crawford FC, Mullan M, Mortimer JA. Is *APOE* -ε4 a risk factor for cognitive impairment in normal aging? Neurology 2000;54:2082–8. 10.1212/WNL.54.11.2082.

[25] Bickeböller H, Campion D, Brice A, Amouyel P, Hannequin D, Didierjean O, et al. Apolipoprotein E and Alzheimer disease: genotype-specific risks by age and sex. Am J Hum Genet 1997;60:439–46.

[26] Neu SC, Pa J, Kukull W, Beekly D, Kuzma A, Gangadharan P, et al. Apolipoprotein E Genotype and Sex Risk Factors for Alzheimer Disease: A Meta-analysis. JAMA Neurol 2017;74:1178–89. 10.1001/jamaneurol.2017.2188.

[27] Hsiung G-YR. Apolipoprotein E 4 genotype as a risk factor for cognitive decline and dementia: data from the Canadian Study of Health and Aging. Canadian Medical Association Journal 2004;171:863–7. 10.1503/cmaj.1031789.

[28] Lambert JC, Ibrahim-Verbaas CA, Harold D, Naj AC, Sims R, Bellenguez C, et al. Meta-analysis of 74,046 individuals identifies 11 new susceptibility loci for Alzheimer’s disease. Nat Genet 2013;45:1452–8. 10.1038/ng.2802.

[29] Jansen IE, Savage JE, Watanabe K, Bryois J, Williams DM, Steinberg S, et al. Genome-wide meta-analysis identifies new loci and functional pathways influencing Alzheimer’s disease risk. Nat Genet 2019;51:404–13. 10.1038/s41588-018-0311-9.

[30] Kunkle BW, Grenier-Boley B, Sims R, Bis JC, Damotte V, Naj AC, et al. Genetic meta-analysis of diagnosed Alzheimer’s disease identifies new risk loci and implicates Aβ, tau, immunity and lipid processing. Nat Genet 2019;51:414–30. 10.1038/s41588-019-0358-2.

[31] Wightman DP, Jansen IE, Savage JE, Shadrin AA, Bahrami S, Holland D, et al. A genome-wide association study with 1,126,563 individuals identifies new risk loci for Alzheimer’s disease. Nat Genet 2021;53:1276–82. 10.1038/s41588-021-00921-z.

[32] Bellenguez C, Küçükali F, Jansen IE, Kleineidam L, Moreno-Grau S, Amin N, et al. New insights into the genetic etiology of Alzheimer’s disease and related dementias. Nat Genet 2022;54:412–36. 10.1038/s41588-022-01024-z.

[33] Martin AR, Gignoux CR, Walters RK, Wojcik GL, Neale BM, Gravel S, et al. Human Demographic History Impacts Genetic Risk Prediction across Diverse Populations. The American Journal of Human Genetics 2017;100:635–49. 10.1016/j.ajhg.2017.03.004.

[34] Naslavsky MS, Suemoto CK, Brito LA, Scliar MO, Ferretti-Rebustini RE, Rodriguez RD, et al. Global and local ancestry modulate APOE association with Alzheimer’s neuropathology and cognitive outcomes in an admixed sample. Mol Psychiatry 2022;27:4800–8. 10.1038/s41380-022-01729-x.

[35] Ganguli M, Chandra V, Kamboh MI, Johnston JM, Dodge HH, Thelma BK, et al. Apolipoprotein E polymorphism and Alzheimer disease: The Indo-US Cross-National Dementia Study. Arch Neurol 2000;57:824–30. 10.1001/archneur.57.6.824.

[36] Agarwal R, Tripathi CB. Association of Apolipoprotein E Genetic Variation in Alzheimer’s Disease in Indian Population: A Meta-Analysis. Am J Alzheimers Dis Other Demen 2014;29:575–82. 10.1177/1533317514531443.

[37] Shankarappa BM, Kota LN, Purushottam M, Nagpal K, Mukherjee O, Viswanath B, et al. Effect of CLU and PICALM polymorphisms on AD risk: A study from south India. Asian Journal of Psychiatry 2017;27:7–11. 10.1016/j.ajp.2016.12.017.

[38] Misra A, Chakrabarti SS, Gambhir IS, Kaur U, Prasad S. APOE4 allele in north Indian elderly patients with dementia or late onset depression-a multiple-disease case control study. Mol Biol Res Commun 2019;8:135–40. 10.22099/mbrc.2019.34417.1427.

[39] Arokiasamy P, Bloom D, Lee J, Feeney K, Ozolins M. Longitudinal aging study in India: Vision, design, implementation, and preliminary findings. Aging in Asia: Findings from new and emerging data initiatives, National Academies Press (US); 2012.

[40] Lee J, Petrosyan S, Khobragade P, Banerjee J, Chien S, Weerman B, et al. Deep phenotyping and genomic data from a nationally representative study on dementia in India. Sci Data 2023;10:45. 10.1038/s41597-023-01941-6.

[41] Leung YY, Valladares O, Chou Y-F, Lin H-J, Kuzma AB, Cantwell L, et al. VCPA: genomic variant calling pipeline and data management tool for Alzheimer’s Disease Sequencing Project. Bioinformatics 2019;35:1768–70. 10.1093/bioinformatics/bty894.

[42] Lee J, Dey AB. Introduction to LASI-DAD: The Longitudinal Aging Study in India-Diagnostic Assessment of Dementia. J American Geriatrics Society 2020;68. 10.1111/jgs.16740.

[43] Morgulis A, Gertz EM, Schäffer AA, Agarwala R. A Fast and Symmetric DUST Implementation to Mask Low-Complexity DNA Sequences. Journal of Computational Biology 2006;13:1028–40. 10.1089/cmb.2006.13.1028.

[44] Conomos MP, Miller MB, Thornton TA. Robust inference of population structure for ancestry prediction and correction of stratification in the presence of relatedness. Genetic Epidemiology 2015;39:276–93.

[45] Ganguli M, Ratcliff G, Chandra V, Sharma S, Gilby J, Pandav R, et al. A Hindi version of the MMSE: the development of a cognitive screening instrument for a largely illiterate rural elderly population in India. International Journal of Geriatric Psychiatry 1995;10:367–77.

[46] Gross AL, Khobragade PY, Meijer E, Saxton JA. Measurement and Structure of Cognition in the Longitudinal Aging Study in India–Diagnostic Assessment of Dementia. J Am Geriatr Soc 2020;68:S11–9. 10.1111/jgs.16738.

[47] NSO. Elderly in India 2021. National Statistical Office, Ministry of Statistics and Programme Implementation, Government of India, New Delhi; 2021.

[48] Chen S, Francioli LC, Goodrich JK, Collins RL, Kanai M, Wang Q, et al. A genome-wide mutational constraint map quantified from variation in 76,156 human genomes. Genetics; 2022. 10.1101/2022.03.20.485034.

[49] Small BJ, Rosnick CB, Fratiglioni L, Bäckman L. Apolipoprotein E and cognitive performance: a meta-analysis. Psychol Aging 2004;19:592–600. 10.1037/0882-7974.19.4.592.

[50] Smith JA, Zhao W, Yu M, Rumfelt KE, Moorjani P, Ganna A, et al. Association Between Episodic Memory and Genetic Risk Factors for Alzheimer’s Disease in South Asians from the Longitudinal Aging Study in India-Diagnostic Assessment of Dementia (LASI-DAD). J Am Geriatr Soc 2020;68 Suppl 3:S45–53. 10.1111/jgs.16735.

[51] Zhao W, Smith JA, Wang YZ, Chintalapati M, Ammous F, Yu M, et al. Polygenic Risk Scores for Alzheimer’s Disease and General Cognitive Function Are Associated With Measures of Cognition in Older South Asians. The Journals of Gerontology: Series A 2023;78:743–52. 10.1093/gerona/glad057.

[52] Davies G, Armstrong N, Bis JC, Bressler J, Chouraki V, Giddaluru S, et al. Genetic contributions to variation in general cognitive function: a meta-analysis of genome-wide association studies in the CHARGE consortium (N=53949). Mol Psychiatry 2015;20:183–92. 10.1038/mp.2014.188.

[53] Laukka EJ, Lövdén M, Herlitz A, Karlsson S, Ferencz B, Pantzar A, et al. Genetic effects on old-age cognitive functioning: A population-based study. Psychology and Aging 2013;28:262–74. 10.1037/a0030829.

[54] Lindenberger U, Nagel IE, Chicherio C, Li S-C, Heekeren HR, Bäckman L. Age-related decline in brain resources modulates genetic effects on cognitive functioning. Front Neurosci 2008;2:234–44. 10.3389/neuro.01.039.2008.

[55] Papenberg G, Salami A, Persson J, Lindenberger U, Bäckman L. Genetics and functional imaging: effects of APOE, BDNF, COMT, and KIBRA in aging. Neuropsychol Rev 2015;25:47–62. 10.1007/s11065-015-9279-8.

[56] Bunce D, Bielak AAM, Anstey KJ, Cherbuin N, Batterham PJ, Easteal S. APOE genotype and cognitive change in young, middle-aged, and older adults living in the community. J Gerontol A Biol Sci Med Sci 2014;69:379–86. 10.1093/gerona/glt103.

[57] Poirier J, Bertrand P, Poirier J, Kogan S, Gauthier S, Poirier J, et al. Apolipoprotein E polymorphism and Alzheimer’s disease. The Lancet 1993;342:697–9. 10.1016/0140-6736(93)91705-Q.

[58] Payami H, Montee KR, Kaye JA, Bird TD, Yu C-E, Wijsman EM, et al. Alzheimer’s disease, apolipoprotein E4, and gender. Jama 1994;271:1316–7.

[59] Hohman TJ, Dumitrescu L, Barnes LL, Thambisetty M, Beecham G, Kunkle B, et al. Sex-Specific Association of Apolipoprotein E With Cerebrospinal Fluid Levels of Tau. JAMA Neurol 2018;75:989. 10.1001/jamaneurol.2018.0821.

[60] Damoiseaux JS, Seeley WW, Zhou J, Shirer WR, Coppola G, Karydas A, et al. Gender Modulates the APOE 4 Effect in Healthy Older Adults: Convergent Evidence from Functional Brain Connectivity and Spinal Fluid Tau Levels. Journal of Neuroscience 2012;32:8254–62. 10.1523/JNEUROSCI.0305-12.2012.

[61] Sampedro F, Vilaplana E, de Leon MJ, Alcolea D, Pegueroles J, Montal V, et al. APOE-by-sex interactions on brain structure and metabolism in healthy elderly controls. Oncotarget 2015;6:26663–74. 10.18632/oncotarget.5185.

[62] Hyman BT, Gomez-Isla T, Briggs M, Chung H, Nichols S, Kohout F, et al. Apolipoprotein E and cognitive change in an elderly population. Ann Neurol 1996;40:55–66. 10.1002/ana.410400111.

[63] Wang X, Zhou W, Ye T, Lin X, Zhang J, for Alzheimer’s Disease Neuroimaging Initiative. Sex Difference in the Association of APOE4 with Memory Decline in Mild Cognitive Impairment. Journal of Alzheimer’s Disease 2019;69:1161–9. 10.3233/JAD-181234.

[64] Gamache J, Yun Y, Chiba-Falek O. Sex-dependent effect of *APOE* on Alzheimer’s disease and other age-related neurodegenerative disorders. Disease Models & Mechanisms 2020;13:dmm045211. 10.1242/dmm.045211.

[65] Lyall DM, Celis-Morales C, Lyall LM, Graham C, Graham N, Mackay DF, et al. Assessing for interaction between *APOE* ε4, sex, and lifestyle on cognitive abilities. Neurology 2019;92:e2691–8. 10.1212/WNL.0000000000007551.

[66] Lee M, Hughes TM, George KM, Griswold ME, Sedaghat S, Simino J, et al. Education and Cardiovascular Health as Effect Modifiers of APOE ε4 on Dementia: The Atherosclerosis Risk in Communities Study. The Journals of Gerontology: Series A 2022;77:1199–207. 10.1093/gerona/glab299.

[67] Lancaster C, Forster S, Tabet N, Rusted J. Putting attention in the spotlight: The influence of APOE genotype on visual search in mid adulthood. Behavioural Brain Research 2017;334:97–104. 10.1016/j.bbr.2017.07.015.

[68] Chey J, Kim J-W, Cho H-Y. Effects of apolipoprotein E phenotypes on the neuropsychological functions of community-dwelling elderly individuals without dementia. Neuroscience Letters 2000;289:230–4. 10.1016/S0304-3940(00)01288-X.

[69] Mostafavi H, Berisa T, Day FR, Perry JRB, Przeworski M, Pickrell JK. Identifying genetic variants that affect viability in large cohorts. PLoS Biol 2017;15:e2002458. 10.1371/journal.pbio.2002458.

[70] McKay GJ, Silvestri G, Chakravarthy U, Dasari S, Fritsche LG, Weber BH, et al. Variations in Apolipoprotein E Frequency With Age in a Pooled Analysis of a Large Group of Older People. American Journal of Epidemiology 2011;173:1357–64. 10.1093/aje/kwr015.

