## Supplemental Material for "Effect of *APOE* ε4 and its modification by sociodemographic characteristics on cognitive measures in South Asians from LASI-DAD"

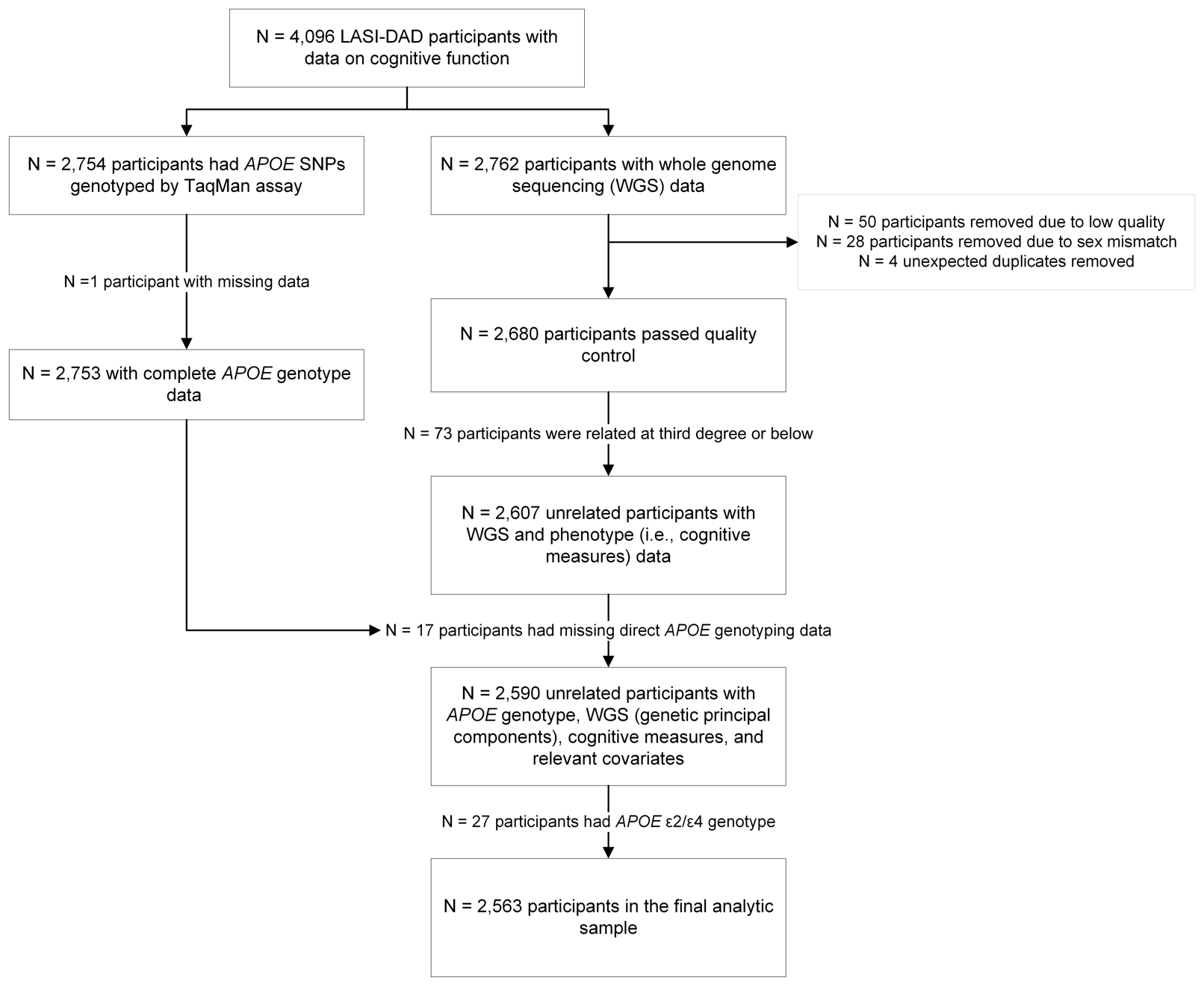

Supplementary Figure 1. Study flow chart.

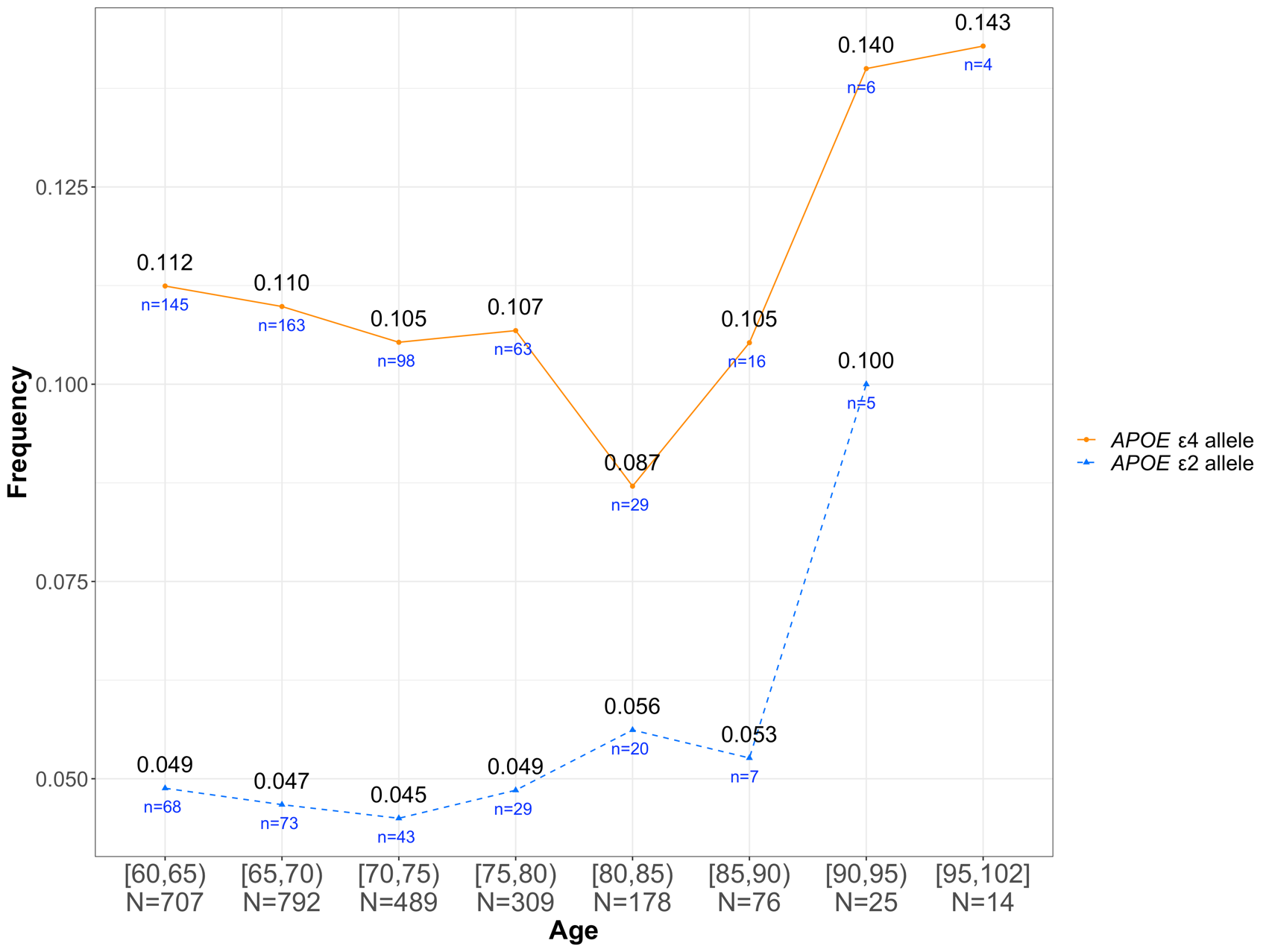

Supplementary Figure 2. Allele frequency of *APOE* ε2 and ε4 by 5-year age group. *APOE* ε4 frequencies are indicated by orange dots and connected by solid orange lines, while *APOE* ε2 frequencies are indicated with blue triangles and connected by dashed blue lines. The numbers under each curve represent the number of participants carrying the respective allele within each age group.

Supplementary Table 1. Allele frequencies of *APOE* ε2, ε3, and ε4 in the full study sample, and by 5-year age group ranging from 60 years to ≥85 years.

|  | **Total sample (N = 2,590)** | **[60,65) (n = 707)** | **[65,70) (n = 792)** | **[70,75) (n =489)** | **[75,80) (n = 309)** | **[80,85) (n = 178)** | **≥85 (n = 115)** | **P value** |
| --- | --- | --- | --- | --- | --- | --- | --- | --- |
| **Allele frequencies** | | | | | | | |  |
| ε2 | 0.048 | 0.049 | 0.047 | 0.045 | 0.049 | 0.056 | 0.057 | 0.950^‡^ |
| ε3 | 0.844 | 0.839 | 0.843 | 0.850 | 0.845 | 0.857 | 0.826 | 0.909^‡^ |
| ε4 | 0.108 | 0.112 | 0.110 | 0.105 | 0.107 | 0.087 | 0.117 | 0.813^‡^ |
| ***APOE* genotype (n, %)** | | | | | | | | 0.904^†^ |
| ε2/ε2 | 5 (0.2%) | 1 (0.1%) | 1 (0.1%) | 1 (0.2%) | 1 (0.3%) | 0 | 1 (0.9%) |  |
| ε2/ε3 | 213 (8.2%) | 57 (8.1%) | 63 (8.0%) | 41 (8.4%) | 23 (7.4%) | 19 (10.7%) | 10 (8.7%) |  |
| ε2/ε4 | 27 (1.0%) | 10 (1.4%) | 9 (1.1%) | 1 (0.2%) | 5 (1.6%) | 1 (0.6%) | 1 (0.9%) |  |
| ε3/ε3 | 1848 (71.4%) | 504 (71.3%) | 565 (71.3%) | 349 (71.4%) | 222 (71.8%) | 130 (73.0%) | 78 (67.8%) |  |
| ε3/ε4 | 461 (17.8%) | 121 (17.1%) | 143 (18.1%) | 92 (18.8%) | 55 (17.8%) | 26 (14.6%) | 24 (20.9%) |  |
| ε4/ε4 | 36 (1.4%) | 14 (2.0%) | 11 (1.4%) | 5 (1.0%) | 3 (1.0%) | 2 (1.1%) | 1 (0.9%) |  |
| ***APOE* ε4 carrier status (n, %)** | | | | | | | | 0.817^†^ |
| ε4 carrier | 524 (20.2%) | 145 (20.5%) | 163 (20.6%) | 98 (20.0%) | 63 (20.4%) | 29 (16.3%) | 26 (22.6%) |  |
| ε4 noncarrier | 2,066 (79.8%) | 562 (79.5%) | 629 (79.4%) | 391 (80.0%) | 246 (79.6%) | 149 (83.7%) | 89 (77.4%) |  |
| ***APOE* ε4 carrier status after removing the *APOE* ε2/ε4 genotype (n, %)** | | | | | | | | 0.843^†^ |
| ε4 carrier | 497 (19.2%) | 135 (19.1%) | 154 (19.4%) | 97 (19.8%) | 58 (18.8%) | 28 (15.7%) | 25 (21.7%) |  |
| ε4 noncarrier | 2,066 (79.8%) | 562 (79.5%) | 629 (79.4%) | 391 (80.0%) | 246 (79.6%) | 149 (83.7%) | 89 (77.4%) |  |

Abbreviations: APOE = apolipoprotein E; LASI-DAD = Diagnostic Assessment of Dementia for the Longitudinal Aging Study of India; HMSE = Hindi Mental State Examination.

*Individuals with the *APOE* ε2/ε4 genotype were not included.

^†^ p-value calculated from chi-square test across the 5-year age groups.

^‡^ p-value calculated from chi-square test for equality of proportions among the 5-year age groups.

Supplementary Table 2. Correlation among cognitive domain scores in LASI-DAD.

|  | HMSE score | General cognitive function | Executive function | Orientation | Language/ fluency | Memory | Visuospatial function |
| --- | --- | --- | --- | --- | --- | --- | --- |
| HMSE score | 1.000 |  |  |  |  |  |  |
| General cognitive function | 0.867 | 1.000 |  |  |  |  |  |
| Executive function | 0.789 | 0.948 | 1.000 |  |  |  |  |
| Orientation | 0.856 | 0.841 | 0.733 | 1.000 |  |  |  |
| Language fluency | 0.773 | 0.816 | 0.713 | 0.655 | 1.000 |  |  |
| Memory | 0.667 | 0.803 | 0.684 | 0.610 | 0.632 | 1.000 |  |
| Visuospatial function | 0.560 | 0.733 | 0.673 | 0.542 | 0.491 | 0.493 | 1.000 |

Abbreviations: LASI-DAD, Longitudinal Aging Study in India-Diagnostic Assessment of Dementia; HMSE = Hindi Mental State Examination.

Supplementary Table 3. Associations of *APOE* ε4 and sociodemographic characteristics with cognitive measures (Model 1).

| **Model 1 (n = 2,563)** | **HMSE score** | | **General cognitive function** | | **Executive function** | | **Orientation** | | **Language/fluency** | | **Memory** | | **Visuospatial function** | |
| --- | --- | --- | --- | --- | --- | --- | --- | --- | --- | --- | --- | --- | --- | --- |
|  | **Beta** | **P value** | **Beta** | **P value** | **Beta** | **P value** | **Beta** | **P value** | **Beta** | **P value** | **Beta** | **P value** | **Beta** | **P value** |
| *APOE* ε4 | -0.978 | 2.44E-05 | -0.142 | 1.69E-04 | -0.117 | 0.002 | -0.141 | 2.86E-05 | -0.095 | 0.005 | -0.134 | 0.001 | -0.063 | 0.097 |
| Age | -0.184 | 1.39E-46 | -0.034 | 1.22E-58 | -0.030 | 9.93E-48 | -0.023 | 1.45E-34 | -0.022 | 4.73E-32 | -0.036 | 7.72E-57 | -0.023 | 9.13E-28 |
| Male | 3.342 | 1.30E-69 | 0.634 | 1.16E-91 | 0.641 | 1.33E-94 | 0.654 | 1.54E-118 | 0.384 | 1.01E-44 | 0.208 | 1.94E-10 | 0.452 | 6.52E-49 |
| Bihar | 3.024 | 1.91E-05 | 0.562 | 1.09E-06 | 0.751 | 6.35E-11 | 0.347 | 0.001 | 0.273 | 0.008 | 0.065 | 0.601 | 0.521 | 6.80E-06 |
| Assam | 3.809 | 4.87E-05 | 0.930 | 1.29E-09 | 1.045 | 7.16E-12 | 0.404 | 0.003 | 0.906 | 4.22E-11 | 0.066 | 0.690 | 0.971 | 2.70E-10 |
| West Bengal | 4.445 | 1.82E-10 | 0.934 | 2.51E-16 | 1.120 | 6.62E-23 | 0.368 | 2.76E-04 | 0.810 | 2.19E-15 | 0.507 | 3.91E-05 | 0.669 | 4.54E-09 |
| Orissa | 1.820 | 0.009 | 0.697 | 1.21E-09 | 0.896 | 4.27E-15 | 0.267 | 0.009 | 0.182 | 0.076 | 0.134 | 0.280 | 0.950 | 1.92E-16 |
| Madhya Pradesh | 2.097 | 0.007 | 0.332 | 0.008 | 0.573 | 4.69E-06 | 0.237 | 0.035 | 0.066 | 0.556 | -0.352 | 0.010 | 0.472 | 1.87E-04 |
| Gujarat | 3.334 | 3.13E-06 | 0.885 | 3.85E-14 | 0.995 | 1.29E-17 | 0.551 | 1.28E-07 | 0.555 | 1.13E-07 | 0.200 | 0.113 | 0.916 | 6.45E-15 |
| Maharashtra | 6.490 | 7.21E-19 | 1.480 | 7.12E-35 | 1.705 | 8.53E-46 | 0.841 | 2.65E-15 | 0.782 | 2.27E-13 | 0.783 | 1.35E-09 | 1.226 | 1.75E-24 |
| Karnataka | 6.346 | 5.69E-13 | 1.358 | 4.24E-21 | 1.578 | 4.55E-28 | 0.598 | 2.91E-06 | 1.122 | 3.32E-18 | 0.794 | 3.42E-07 | 0.949 | 4.44E-11 |
| Punjab | 5.034 | 1.25E-12 | 0.879 | 2.95E-14 | 0.883 | 1.60E-14 | 0.670 | 8.54E-11 | 0.645 | 4.57E-10 | 0.557 | 8.85E-06 | 0.678 | 4.83E-09 |
| Kerala | 8.726 | 2.50E-22 | 1.829 | 2.02E-35 | 1.897 | 2.56E-38 | 1.010 | 9.44E-15 | 1.462 | 1.07E-28 | 1.297 | 3.03E-16 | 1.176 | 1.01E-15 |
| Tamil Nadu | 7.068 | 3.12E-15 | 1.601 | 1.22E-27 | 1.781 | 5.51E-34 | 0.778 | 2.33E-09 | 1.338 | 2.67E-24 | 0.968 | 1.02E-09 | 1.130 | 1.33E-14 |
| Telangana | 3.480 | 1.56E-05 | 0.999 | 3.60E-14 | 1.157 | 1.19E-18 | 0.364 | 0.002 | 0.702 | 2.67E-09 | 0.488 | 0.001 | 1.012 | 2.08E-14 |
| Uttranchal | 2.339 | 0.003 | 0.548 | 2.72E-05 | 0.696 | 8.53E-08 | 0.318 | 0.006 | 0.343 | 0.003 | -0.045 | 0.748 | 0.595 | 5.71E-06 |
| Haryana | 1.873 | 0.003 | 0.326 | 0.002 | 0.522 | 4.28E-07 | 0.209 | 0.024 | 0.132 | 0.156 | -0.195 | 0.084 | 0.344 | 0.001 |
| Delhi | 3.093 | 5.29E-06 | 0.820 | 1.63E-13 | 1.042 | 5.21E-21 | 0.544 | 4.06E-08 | 0.458 | 3.90E-06 | 0.246 | 0.041 | 0.508 | 4.99E-06 |
| Rajasthan | 2.205 | 0.001 | 0.400 | 2.62E-04 | 0.646 | 3.28E-09 | 0.192 | 0.050 | 0.453 | 4.15E-06 | -0.397 | 0.001 | 0.296 | 0.007 |
| Uttar Pradesh | 3.388 | 2.73E-07 | 0.532 | 7.30E-07 | 0.719 | 1.80E-11 | 0.433 | 6.23E-06 | 0.355 | 2.25E-04 | -0.162 | 0.164 | 0.516 | 1.71E-06 |
| PC1 | 67.706 | 2.59E-18 | 14.069 | 1.71E-28 | 13.618 | 3.90E-27 | 9.039 | 1.16E-15 | 9.919 | 2.14E-18 | 12.653 | 3.72E-20 | 8.605 | 1.06E-11 |
| PC2 | 29.839 | 0.001 | 7.682 | 5.42E-07 | 8.013 | 1.46E-07 | 4.169 | 0.002 | 3.531 | 0.010 | 6.819 | 4.22E-05 | 6.589 | 1.83E-05 |
| PC3 | 12.212 | 0.155 | 2.670 | 0.057 | 2.470 | 0.076 | 1.528 | 0.222 | 0.704 | 0.575 | 2.860 | 0.060 | 3.004 | 0.033 |
| PC4 | 27.901 | 1.71E-05 | 5.421 | 3.08E-07 | 4.967 | 2.35E-06 | 3.485 | 2.26E-04 | 2.411 | 0.011 | 6.086 | 1.25E-07 | 4.531 | 2.00E-05 |
| PC5 | 16.919 | 0.001 | 3.779 | 1.25E-05 | 4.163 | 1.30E-06 | 2.390 | 0.002 | 1.090 | 0.159 | 3.244 | 0.001 | 3.339 | 1.20E-04 |
| PC6 | 5.613 | 0.281 | 0.453 | 0.594 | 0.061 | 0.942 | 0.697 | 0.358 | 1.426 | 0.061 | 0.072 | 0.938 | -0.534 | 0.531 |
| PC7 | 4.376 | 0.362 | 0.770 | 0.325 | 1.312 | 0.092 | 0.321 | 0.646 | 0.104 | 0.882 | 0.541 | 0.525 | 0.385 | 0.624 |
| PC8 | -5.569 | 0.286 | -0.736 | 0.388 | -1.261 | 0.137 | -0.470 | 0.537 | -0.341 | 0.655 | 0.594 | 0.521 | -0.589 | 0.491 |
| PC9 | -10.755 | 0.070 | -1.983 | 0.040 | -1.543 | 0.109 | -2.224 | 0.010 | -1.988 | 0.022 | -1.001 | 0.341 | -1.021 | 0.293 |
| PC10 | 1.528 | 0.760 | 0.073 | 0.929 | 0.379 | 0.640 | -0.125 | 0.863 | 0.080 | 0.913 | 0.239 | 0.787 | -0.562 | 0.492 |
| **Total R^2^** | **0.281** | | **0.339** | | **0.328** | | **0.299** | | **0.307** | | **0.252** | | **0.186** | |

Abbreviations: APOE = apolipoprotein E; PC = principal component; HMSE = Hindi Mental State Examination.

Supplementary Table 4. Associations of *APOE* ε4 and sociodemographic characteristics with cognitive measures (Model 2).

| **Model 2 (n = 2,548)** | **HMSE score** | | **General cognitive function** | | **Executive function** | | **Orientation** | | **Language/ fluency** | | **Memory** | | **Visuospatial function** | |
| --- | --- | --- | --- | --- | --- | --- | --- | --- | --- | --- | --- | --- | --- | --- |
|  | **Beta** | **P value** | **Beta** | **P value** | **Beta** | **P value** | **Beta** | **P value** | **Beta** | **P value** | **Beta** | **P value** | **Beta** | **P value** |
| *APOE* ε4 | -0.710 | 0.001 | -0.079 | 0.006 | -0.057 | 0.054 | -0.096 | 0.001 | -0.052 | 0.064 | -0.087 | 0.019 | -0.019 | 0.576 |
| Age | -0.154 | 1.98E-39 | -0.026 | 4.47E-58 | -0.022 | 3.18E-41 | -0.018 | 5.50E-28 | -0.017 | 4.00E-26 | -0.031 | 6.72E-49 | -0.016 | 7.29E-19 |
| Male | 1.813 | 2.91E-23 | 0.249 | 2.11E-23 | 0.286 | 4.05E-28 | 0.398 | 2.62E-53 | 0.092 | 1.62E-04 | -0.068 | 0.034 | 0.154 | 8.65E-08 |
| Bihar | 2.096 | 0.001 | 0.305 | 0.001 | 0.523 | 2.23E-08 | 0.180 | 0.050 | 0.033 | 0.704 | -0.103 | 0.372 | 0.330 | 0.001 |
| Assam | 1.008 | 0.247 | 0.290 | 0.015 | 0.462 | 1.97E-04 | -0.066 | 0.586 | 0.352 | 0.003 | -0.332 | 0.031 | 0.532 | 1.21E-04 |
| West Bengal | 1.691 | 0.009 | 0.287 | 0.001 | 0.519 | 2.35E-08 | -0.092 | 0.315 | 0.288 | 0.001 | 0.054 | 0.640 | 0.244 | 0.018 |
| Orissa | -1.026 | 0.119 | 0.031 | 0.730 | 0.292 | 0.002 | -0.216 | 0.019 | -0.390 | 1.10E-05 | -0.316 | 0.007 | 0.501 | 1.77E-06 |
| Madhya Pradesh | 1.344 | 0.058 | 0.123 | 0.206 | 0.385 | 1.38E-04 | 0.101 | 0.310 | -0.114 | 0.232 | -0.505 | 5.64E-05 | 0.331 | 0.003 |
| Gujarat | 0.785 | 0.235 | 0.252 | 0.005 | 0.416 | 1.00E-05 | 0.102 | 0.273 | 0.057 | 0.520 | -0.263 | 0.024 | 0.465 | 1.00E-05 |
| Maharashtra | 3.202 | 2.97E-06 | 0.660 | 2.40E-12 | 0.934 | 1.83E-21 | 0.276 | 0.004 | 0.176 | 0.056 | 0.180 | 0.135 | 0.633 | 6.42E-09 |
| Karnataka | 3.388 | 3.04E-05 | 0.647 | 6.44E-09 | 0.912 | 4.08E-15 | 0.097 | 0.394 | 0.594 | 5.99E-08 | 0.268 | 0.061 | 0.450 | 4.81E-04 |
| Punjab | 4.009 | 7.28E-10 | 0.589 | 3.96E-11 | 0.626 | 1.39E-11 | 0.481 | 1.31E-07 | 0.406 | 3.41E-06 | 0.343 | 0.003 | 0.453 | 1.14E-05 |
| Kerala | 3.894 | 3.16E-06 | 0.654 | 1.14E-08 | 0.799 | 2.07E-11 | 0.195 | 0.095 | 0.580 | 2.64E-07 | 0.442 | 0.003 | 0.331 | 0.012 |
| Tamil Nadu | 3.672 | 1.05E-05 | 0.751 | 5.15E-11 | 0.982 | 1.69E-16 | 0.198 | 0.090 | 0.695 | 6.36E-10 | 0.341 | 0.020 | 0.540 | 4.50E-05 |
| Telangana | 0.948 | 0.203 | 0.375 | 2.39E-04 | 0.574 | 6.46E-08 | -0.069 | 0.508 | 0.231 | 0.021 | 0.022 | 0.866 | 0.586 | 7.64E-07 |
| Uttranchal | 0.809 | 0.269 | 0.159 | 0.113 | 0.347 | 8.66E-04 | 0.052 | 0.609 | 0.011 | 0.914 | -0.318 | 0.014 | 0.323 | 0.005 |
| Haryana | 1.026 | 0.078 | 0.091 | 0.256 | 0.310 | 1.86E-04 | 0.054 | 0.511 | -0.060 | 0.444 | -0.365 | 3.94E-04 | 0.166 | 0.073 |
| Delhi | 0.830 | 0.199 | 0.224 | 0.011 | 0.477 | 2.31E-07 | 0.142 | 0.118 | 0.018 | 0.832 | -0.196 | 0.086 | 0.096 | 0.351 |
| Rajasthan | 1.235 | 0.047 | 0.125 | 0.142 | 0.407 | 4.25E-06 | 0.008 | 0.925 | 0.224 | 0.008 | -0.599 | 5.18E-08 | 0.075 | 0.450 |
| Uttar Pradesh | 2.046 | 0.001 | 0.145 | 0.084 | 0.351 | 5.85E-05 | 0.194 | 0.024 | 0.074 | 0.369 | -0.453 | 2.95E-05 | 0.231 | 0.018 |
| PC1 | 17.924 | 0.026 | 3.970 | 3.26E-04 | 4.007 | 4.77E-04 | 1.118 | 0.322 | 2.134 | 0.049 | 5.777 | 4.96E-05 | 2.491 | 0.052 |
| PC2 | 13.650 | 0.117 | 3.614 | 0.002 | 4.045 | 0.001 | 1.495 | 0.220 | 1.219 | 0.299 | 3.462 | 0.024 | 3.253 | 0.019 |
| PC3 | 13.905 | 0.083 | 2.722 | 0.013 | 2.473 | 0.030 | 1.797 | 0.110 | 1.094 | 0.312 | 2.613 | 0.065 | 2.878 | 0.024 |
| PC4 | 10.959 | 0.074 | 1.903 | 0.023 | 1.576 | 0.071 | 0.687 | 0.424 | 0.006 | 0.994 | 3.544 | 0.001 | 2.249 | 0.021 |
| PC5 | 10.217 | 0.036 | 2.247 | 0.001 | 2.638 | 1.44E-04 | 1.299 | 0.057 | -0.046 | 0.944 | 2.154 | 0.012 | 2.467 | 0.001 |
| PC6 | 1.948 | 0.681 | -0.250 | 0.700 | -0.608 | 0.367 | 0.113 | 0.865 | 0.927 | 0.146 | -0.473 | 0.572 | -0.879 | 0.242 |
| PC7 | 3.106 | 0.476 | 0.310 | 0.604 | 0.874 | 0.159 | 0.075 | 0.902 | -0.184 | 0.754 | 0.107 | 0.889 | 0.062 | 0.928 |
| PC8 | -1.382 | 0.771 | 0.135 | 0.835 | -0.469 | 0.487 | 0.268 | 0.687 | 0.449 | 0.482 | 1.271 | 0.129 | -0.293 | 0.697 |
| PC9 | -1.752 | 0.746 | 0.187 | 0.801 | 0.382 | 0.620 | -0.648 | 0.393 | -0.277 | 0.704 | 0.681 | 0.477 | 0.566 | 0.510 |
| PC10 | 1.051 | 0.816 | 0.117 | 0.849 | 0.429 | 0.504 | -0.207 | 0.743 | 0.055 | 0.928 | 0.297 | 0.709 | -0.420 | 0.558 |
| Education level |  |  |  |  |  |  |  |  |  |  |  |  |  |  |
| Upper secondary or vocational training | 1.480 | 6.39E-09 | 0.433 | 1.92E-34 | 0.458 | 1.10E-35 | 0.227 | 1.95E-10 | 0.179 | 1.86E-07 | 0.354 | 4.51E-15 | 0.377 | 1.81E-20 |
| Tertiary | 2.392 | 5.79E-07 | 0.843 | 8.10E-37 | 0.893 | 3.11E-38 | 0.361 | 7.24E-08 | 0.327 | 4.02E-07 | 0.787 | 2.12E-20 | 0.583 | 2.05E-14 |
| Cannot read or write | -3.383 | 1.52E-50 | -0.771 | 3.86E-127 | -0.660 | 3.78E-90 | -0.580 | 2.33E-73 | -0.732 | 1.04E-119 | -0.500 | 2.39E-36 | -0.563 | 4.27E-55 |
| Rural village | -0.330 | 0.112 | -0.104 | 2.45E-04 | -0.118 | 6.97E-05 | -0.071 | 0.015 | -0.031 | 0.263 | -0.106 | 0.004 | -0.052 | 0.114 |
| Scheduled caste | -0.937 | 0.003 | -0.092 | 3.32E-02 | -0.101 | 0.024 | -0.138 | 0.002 | -0.085 | 0.046 | -0.012 | 0.834 | -0.020 | 0.683 |
| Scheduled tribe | -1.715 | 0.001 | -0.152 | 3.27E-02 | -0.141 | 0.057 | -0.252 | 0.001 | -0.268 | 0.000 | -0.014 | 0.882 | 0.113 | 0.171 |
| Other backward class (obc) | -0.393 | 0.081 | -0.027 | 3.77E-01 | -0.024 | 0.457 | -0.060 | 0.057 | -0.035 | 0.248 | 0.007 | 0.850 | -0.016 | 0.662 |
| Per capita household consumption |  |  |  |  |  |  |  |  |  |  |  |  |  |  |
| Q2 | 0.353 | 0.179 | 0.054 | 1.31E-01 | 0.063 | 0.091 | 0.043 | 0.244 | 0.007 | 0.835 | 0.024 | 0.603 | 0.036 | 0.383 |
| Q3 | 0.727 | 0.007 | 0.086 | 1.97E-02 | 0.095 | 0.013 | 0.076 | 0.045 | -0.016 | 0.661 | 0.038 | 0.421 | 0.110 | 0.010 |
| Q4 | 0.862 | 0.002 | 0.129 | 6.44E-04 | 0.119 | 0.002 | 0.123 | 0.001 | 0.026 | 0.480 | 0.123 | 0.012 | 0.109 | 0.012 |
| Q5 | 0.544 | 0.057 | 0.106 | 6.58E-03 | 0.099 | 0.015 | 0.078 | 0.053 | 0.036 | 0.345 | 0.119 | 0.018 | 0.060 | 0.183 |
| **Total R^2^** | **0.421** | | **0.624** | | **0.582** | | **0.477** | | **0.525** | | **0.403** | | **0.379** | |

Abbreviations: APOE = apolipoprotein E; PC = principal component; HMSE = Hindi Mental State Examination.

Supplementary Table 5. Two-way interaction between *APOE* ε4 carrier and educational level on cognitive measures.

|  | **Model 1 (n = 2,563)** | | **Model 2 (n = 2,548)** | |
| --- | --- | --- | --- | --- |
|  | **Beta** | **P value** | **Beta** | **P value** |
| **HMSE score** |  |  |  |  |
| *APOE* ε4 | **-0.872** | **4.25E-04** | **-0.770** | **0.001** |
| Upper secondary or vocational training | **3.498** | **7.97E-43** | **1.453** | **7.29E-08** |
| Tertiary | **4.481** | **8.71E-18** | **2.235** | **1.58E-05** |
| *APOE* ε4*Upper secondary or vocational training | 0.312 | 0.588 | 0.160 | 0.772 |
| *APOE* ε4*Tertiary | 0.878 | 0.474 | 0.931 | 0.424 |
| **General cognitive function** |  |  |  |  |
| *APOE* ε4 | **-0.100** | **0.006** | **-0.082** | **0.011** |
| Upper secondary or vocational training | **0.889** | **3.73E-115** | **0.432** | **6.40E-31** |
| Tertiary | **1.334** | **2.51E-64** | **0.827** | **1.01E-30** |
| *APOE* ε4*Upper secondary or vocational training | 0.024 | 0.775 | 0.001 | 0.986 |
| *APOE* ε4*Tertiary | 0.081 | 0.653 | 0.095 | 0.551 |
| **Executive function** |  |  |  |  |
| *APOE* ε4 | **-0.075** | **0.040** | -0.059 | 0.078 |
| Upper secondary or vocational training | **0.855** | **7.44E-107** | **0.460** | **2.46E-32** |
| Tertiary | **1.316** | **1.82E-62** | **0.873** | **1.11E-31** |
| *APOE* ε4*Upper secondary or vocational training | 0.010 | 0.907 | -0.013 | 0.867 |
| *APOE* ε4*Tertiary | 0.109 | 0.550 | 0.121 | 0.465 |
| **Orientation** |  |  |  |  |
| *APOE* ε4 | **-0.110** | **0.002** | **-0.091** | **0.006** |
| Upper secondary or vocational training | **0.585** | **1.09E-56** | **0.236** | **4.18E-10** |
| Tertiary | **0.724** | **5.06E-22** | **0.336** | **3.53E-06** |
| *APOE* ε4*Upper secondary or vocational training | -0.029 | 0.723 | -0.057 | 0.460 |
| *APOE* ε4*Tertiary | 0.141 | 0.423 | 0.150 | 0.359 |
| **Language/fluency** |  |  |  |  |
| *APOE* ε4 | -0.062 | 0.082 | -0.048 | 0.128 |
| Upper secondary or vocational training | **0.605** | **1.54E-60** | **0.187** | **2.76E-07** |
| Tertiary | **0.751** | **1.27E-23** | **0.302** | **1.44E-05** |
| *APOE* ε4*Upper secondary or vocational training | -0.030 | 0.720 | -0.050 | 0.498 |
| *APOE* ε4*Tertiary | 0.137 | 0.437 | 0.146 | 0.351 |
| **Memory** |  |  |  |  |
| *APOE* ε4 | **-0.092** | **0.033** | -0.081 | 0.051 |
| Upper secondary or vocational training | **0.670** | **3.51E-51** | **0.361** | **4.32E-14** |
| Tertiary | **1.132** | **3.79E-35** | **0.775** | **3.33E-17** |
| *APOE* ε4*Upper secondary or vocational training | -0.042 | 0.673 | -0.044 | 0.647 |
| *APOE* ε4*Tertiary | 0.063 | 0.770 | 0.073 | 0.722 |
| **Visuospatial function** |  |  |  |  |
| *APOE* ε4 | -0.056 | 0.156 | -0.048 | 0.195 |
| Upper secondary or vocational training | **0.675** | **3.77E-61** | **0.349** | **4.52E-16** |
| Tertiary | **0.942** | **1.42E-29** | **0.588** | **1.00E-12** |
| *APOE* ε4*Upper secondary or vocational training | **0.183** | **0.046*** | **0.175** | **0.045*** |
| *APOE* ε4*Tertiary | -0.049 | 0.803 | -0.030 | 0.872 |

Abbreviations: APOE = apolipoprotein E; LASI-DAD = Diagnostic Assessment of Dementia for the Longitudinal Aging Study of India; HMSE = Hindi Mental State Examination.

Model 1 adjusted for age, sex (male), state of residence, top 10 genetic PCs, education (upper secondary or vocational training and tertiary education), *APOE* ε4 × upper secondary or vocational training education, and *APOE* ε4 × upper secondary or tertiary education.

Model 2 adjusted for age, sex (male), state of residence, top 10 genetic PCs, education (upper secondary or vocational training and tertiary education), literacy, urban/rural residence, caste, quintiles of per capita household consumption, *APOE* ε4 × upper secondary or vocational training education, and *APOE* ε4 × upper secondary or tertiary education.

Beta coefficient and p-value in bold indicates statistically significant association at p<0.05.

Asterisks (*) indicates significant *APOE* ε4 by education interaction terms.

Supplementary Table 6. Associations between *APOE* ε4 carrier and cognitive measures stratified by education level.

|  | Model 1 | | | | | | Model 2 | | | | | |
| --- | --- | --- | --- | --- | --- | --- | --- | --- | --- | --- | --- | --- |
|  | **Less than lower secondary (n=1,926)** | | **Upper secondary or vocational training (n=542)** | | **Tertiary**  **(n=95)** | | **Less than lower secondary (n=1,914)** | | **Upper secondary or vocational training (n=539)** | | **Tertiary**  **(n=95)** | |
| **Cognitive Measure** | **Beta** | **P value** | **Beta** | **P value** | **Beta** | **P value** | **Beta** | **P value** | **Beta** | **P value** | **Beta** | **P value** |
| **HMSE score** | **-0.861** | **0.001** | -0.744 | 0.050 | -0.025 | 0.963 | **-0.758** | **0.003** | **-0.756** | **0.049** | 0.041 | 0.944 |
| **General cognitive function** | **-0.102** | **0.005** | -0.110 | 0.162 | 0.104 | 0.502 | **-0.082** | **0.008** | -0.110 | 0.156 | 0.120 | 0.471 |
| **Executive function** | **-0.076** | **0.037** | -0.096 | 0.231 | 0.186 | 0.309 | -0.059 | 0.071 | -0.093 | 0.240 | 0.190 | 0.335 |
| **Orientation** | **-0.112** | **0.003** | **-0.149** | **0.014** | 0.002 | 0.978 | **-0.093** | **0.007** | **-0.159** | **0.009** | -0.006 | 0.936 |
| **Language/fluency** | -0.061 | 0.096 | -0.091 | 0.187 | 0.034 | 0.819 | -0.045 | 0.161 | -0.093 | 0.161 | -0.006 | 0.972 |
| **Memory** | **-0.096** | **0.020** | -0.161 | 0.127 | 0.117 | 0.614 | **-0.081** | **0.038** | -0.155 | 0.142 | 0.096 | 0.704 |
| **Visuospatial function** | -0.055 | 0.150 | 0.089 | 0.334 | -0.056 | 0.811 | -0.047 | 0.189 | 0.087 | 0.346 | 0.047 | 0.852 |

Abbreviations: APOE = apolipoprotein E; LASI-DAD = Diagnostic Assessment of Dementia for the Longitudinal Aging Study of India; HMSE = Hindi Mental State Examination.

Model 1 adjusted for age, sex (male), state of residence, top 10 genetic PCs.

Model 2 adjusted for age, sex (male), state of residence, top 10 genetic PCs, literacy, urban/rural residence, caste, and quintiles of per capita household consumption.

Beta coefficient and p-value in bold indicates statistically significant association at p<0.05.

Supplementary Table 7. Two-way interaction between *APOE* ε4 carrier and dichotomous education level on cognitive measures.

|  | **Model 1 (n = 2,563)** | | **Model 2 (n = 2,548)** | |
| --- | --- | --- | --- | --- |
|  | **Beta** | **P value** | **Beta** | **P value** |
| **HMSE score** |  |  |  |  |
| *APOE* ε4 | **-0.986** | **0.001** | **-0.880** | **0.002** |
| > 0 years of education | **4.396** | **4.79E-87** | **2.576** | **4.06E-22** |
| *APOE* ε4 × > 0 years of education | 0.410 | 0.330 | 0.346 | 0.401 |
| **General cognitive function** |  |  |  |  |
| *APOE* ε4 | **-0.121** | **0.008** | **-0.102** | **0.013** |
| > 0 years of education | **0.899** | **2.33E-144** | **0.360** | **2.62E-21** |
| *APOE* ε4 × > 0 years of education | 0.035 | 0.588 | 0.034 | 0.568 |
| **Executive function** |  |  |  |  |
| *APOE* ε4 | -0.083 | 0.070 | -0.066 | 0.122 |
| > 0 years of education | **0.844** | **4.51E-126** | **0.369** | **8.96E-21** |
| *APOE* ε4 × > 0 years of education | 0.004 | 0.956 | 0.003 | 0.965 |
| **Orientation** |  |  |  |  |
| *APOE* ε4 | **-0.114** | **0.007** | **-0.094** | **0.020** |
| > 0 years of education | **0.701** | **2.50E-105** | **0.362** | **3.60E-22** |
| *APOE* ε4 × > 0 years of education | 0.005 | 0.928 | -0.006 | 0.913 |
| **Language/fluency** |  |  |  |  |
| *APOE* ε4 | **-0.091** | **0.033** | -0.076 | 0.052 |
| > 0 years of education | **0.650** | **2.44E-89** | **0.163** | **6.69E-06** |
| *APOE* ε4 × > 0 years of education | 0.051 | 0.403 | 0.045 | 0.429 |
| **Memory** |  |  |  |  |
| *APOE* ε4 | **-0.135** | **0.013** | **-0.126** | **0.016** |
| > 0 years of education | **0.602** | **1.52E-50** | **0.205** | **2.00E-05** |
| *APOE* ε4 × > 0 years of education | 0.056 | 0.475 | 0.067 | 0.373 |
| **Visuospatial function** |  |  |  |  |
| *APOE* ε4 | -0.056 | 0.247 | -0.050 | 0.286 |
| > 0 years of education | **0.692** | **8.06E-80** | **0.319** | **1.36E-13** |
| *APOE* ε4 × > 0 years of education | 0.047 | 0.495 | 0.053 | 0.433 |

Abbreviations: APOE = apolipoprotein E; LASI-DAD = Diagnostic Assessment of Dementia for the Longitudinal Aging Study of India; HMSE = Hindi Mental State Examination.

Model 1 adjusted for age, sex (male), state of residence, top 10 genetic PCs, education (> 0 years of education), and *APOE* ε4 × > 0 years of education.

Model 2 adjusted for age, sex (male), state of residence, top 10 genetic PCs, education (> 0 years of education), literacy, urban/rural residence, caste, quintiles of per capita household consumption, and *APOE* ε4 × > 0 years of education.

Beta coefficient and p-value in bold indicates statistically significant association at p<0.05.

Supplementary Table 8. Associations between *APOE* ε4 carrier and cognitive measures stratified by education (dichotomized at having received some formal schooling).

|  | Model 1 | | | | Model 2 | | | |
| --- | --- | --- | --- | --- | --- | --- | --- | --- |
|  | **0 years (n = 1,249)** | | **>0 years  (n = 1,314)** | | **0 years  (n = 1,239)** | | **>0 years  (n = 1,348)** | |
| **Cognitive Measure** | **Beta** | **P value** | **Beta** | **P value** | **Beta** | **P value** | **Beta** | **P value** |
| **HMSE score** | **-0.902** | **0.005** | **-0.695** | **0.011** | **-0.779** | **0.016** | **-0.630** | **0.016** |
| **General cognitive function** | **-0.122** | **0.002** | -0.100 | 0.056 | **-0.101** | **0.006** | -0.076 | 0.099 |
| **Executive function** | **-0.084** | **0.031** | -0.089 | 0.094 | -0.065 | 0.084 | -0.067 | 0.170 |
| **Orientation** | **-0.114** | **0.008** | **-0.116** | **0.007** | **-0.089** | **0.034** | **-0.105** | **0.009** |
| **Language/fluency** | **-0.088** | **0.035** | -0.048 | 0.298 | -0.070 | 0.085 | -0.036 | 0.368 |
| **Memory** | **-0.141** | **0.003** | -0.096 | 0.120 | **-0.129** | **0.006** | -0.073 | 0.216 |
| **Visuospatial function** | -0.057 | 0.167 | -0.025 | 0.663 | -0.052 | 0.203 | -0.005 | 0.922 |

Abbreviations: APOE = apolipoprotein E; LASI-DAD = Diagnostic Assessment of Dementia for the Longitudinal Aging Study of India; HMSE = Hindi Mental State Examination.

Model 1 adjusted for age, sex (male), state of residence, top 10 genetic PCs.

Model 2 adjusted for age, sex (male), state of residence, top 10 genetic PCs, literacy, urban/rural residence, caste, and quintiles of per capita household consumption.

Beta coefficient and p-value in bold indicates statistically significant association at p<0.05.

Supplementary Table 9. Main effects of *APOE* genotypes on cognitive measures in LASI-DAD

|  | **Model 1 (n = 2,558)** | | **Model 2 (n = 2,543)** | |
| --- | --- | --- | --- | --- |
|  | **Beta** | **P value** | **Beta** | **P value** |
| **HMSE score** |  |  |  |  |
| ε3/ε3 | ref | ref | ref | ref |
| ε2/ε3 | 0.181 | 0.589 | 0.009 | 0.977 |
| ε3/ε4 | **-0.969** | **5.99E-05** | **-0.727** | **0.001** |
| ε4/ε4 | -0.818 | 0.293 | -0.495 | 0.481 |
| **General cognitive function** |  |  |  |  |
| ε3/ε3 | ref | ref | ref | ref |
| ε2/ε3 | 0.006 | 0.916 | -0.039 | 0.349 |
| ε3/ε4 | **-0.140** | **3.61E-04** | **-0.083** | **0.005** |
| ε4/ε4 | -0.150 | 0.237 | -0.079 | 0.414 |
| **Executive function** |  |  |  |  |
| ε3/ε3 | ref | ref | ref | ref |
| ε2/ε3 | 0.001 | 0.989 | -0.038 | 0.381 |
| ε3/ε4 | **-0.113** | **0.004** | -0.060 | 0.055 |
| ε4/ε4 | -0.162 | 0.200 | -0.083 | 0.406 |
| **Orientation** |  |  |  |  |
| ε3/ε3 | ref | ref | ref | ref |
| ε2/ε3 | -0.015 | 0.756 | -0.053 | 0.214 |
| ε3/ε4 | **-0.143** | **4.90E-05** | **-0.102** | **0.001** |
| ε4/ε4 | -0.139 | 0.220 | -0.094 | 0.339 |
| **Language/fluency** |  |  |  |  |
| ε3/ε3 | ref | ref | ref | ref |
| ε2/ε3 | 0.092 | 0.059 | 0.060 | 0.142 |
| ε3/ε4 | **-0.090** | **0.011** | -0.050 | 0.092 |
| ε4/ε4 | -0.018 | 0.871 | -0.001 | 0.992 |
| **Memory** |  |  |  |  |
| ε3/ε3 | ref | ref | ref | ref |
| ε2/ε3 | 0.015 | 0.799 | -0.015 | 0.783 |
| ε3/ε4 | **-0.130** | **0.002** | **-0.088** | **0.023** |
| ε4/ε4 | -0.170 | 0.216 | -0.102 | 0.410 |
| **Visuospatial function** |  |  |  |  |
| ε3/ε3 | ref | ref | ref | ref |
| ε2/ε3 | -0.080 | 0.146 | **-0.119** | **0.014** |
| ε3/ε4 | -0.075 | 0.057 | -0.035 | 0.307 |
| ε4/ε4 | -0.022 | 0.863 | 0.020 | 0.861 |

Abbreviations: APOE = apolipoprotein E; LASI-DAD = Diagnostic Assessment of Dementia for the Longitudinal Aging Study of India; HMSE = Hindi Mental State Examination; ref = referent.

Model 1 adjusted for age, sex (male), state of residence, and top 10 genetic PCs.

Model 2 adjusted for age, sex (male), state of residence, top 10 genetic PCs, education (upper secondary or vocational training and tertiary education), literacy, urban/rural residence, caste, and quintiles of per capita household consumption.

Supplementary Table 10. Two-way interaction between *APOE* genotypes and median age on cognitive measures

|  | **Model 1 (n = 2,558)** | | **Model 2 (n = 2,543)** | |
| --- | --- | --- | --- | --- |
|  | **Beta** | **P value** | **Beta** | **P value** |
| **HMSE score** |  |  |  |  |
| ε3/ε3*Age > 68 | ref | ref | ref | ref |
| ε2/ε3*Age > 68 | -0.234 | 0.732 | -0.270 | 0.663 |
| ε3/ε4*Age > 68 | -0.406 | 0.407 | -0.712 | 0.107 |
| ε4/ε4*Age > 68 | -0.799 | 0.623 | -0.571 | 0.696 |
| **General cognitive function*** |  |  |  |  |
| ε3/ε3*Age > 68 | ref | ref | ref | ref |
| ε2/ε3*Age > 68 | -0.081 | 0.467 | -0.104 | 0.220 |
| ε3/ε4*Age > 68 | -0.042 | 0.603 | **-0.122** | **0.045** |
| ε4/ε4*Age > 68 | -0.384 | 0.147 | -0.332 | 0.098 |
| **Executive function** |  |  |  |  |
| ε3/ε3*Age > 68 | ref | ref | ref | ref |
| ε2/ε3*Age > 68 | -0.068 | 0.540 | -0.085 | 0.332 |
| ε3/ε4*Age > 68 | -0.016 | 0.843 | -0.098 | 0.120 |
| ε4/ε4*Age > 68 | -0.358 | 0.174 | -0.326 | 0.116 |
| **Orientation*** |  |  |  |  |
| ε3/ε3*Age > 68 | ref | ref | ref | ref |
| ε2/ε3*Age > 68 | -0.037 | 0.708 | -0.067 | 0.435 |
| ε3/ε4*Age > 68 | -0.082 | 0.247 | **-0.128** | **0.037** |
| ε4/ε4*Age > 68 | -0.170 | 0.469 | -0.123 | 0.545 |
| **Language/fluency*** |  |  |  |  |
| ε3/ε3*Age > 68 | ref | ref | ref | ref |
| ε2/ε3*Age > 68 | 0.003 | 0.977 | -0.012 | 0.883 |
| ε3/ε4*Age > 68 | -0.088 | 0.216 | **-0.133** | **0.025** |
| ε4/ε4*Age > 68 | -0.321 | 0.173 | -0.261 | 0.182 |
| **Memory** |  |  |  |  |
| ε3/ε3*Age > 68 | ref | ref | ref | ref |
| ε2/ε3*Age > 68 | -0.123 | 0.308 | -0.147 | 0.180 |
| ε3/ε4*Age > 68 | -0.052 | 0.551 | -0.115 | 0.139 |
| ε4/ε4*Age > 68 | -0.286 | 0.319 | -0.255 | 0.322 |
| **Visuospatial function** |  |  |  |  |
| ε3/ε3*Age > 68 | ref | ref | ref | ref |
| ε2/ε3*Age > 68 | -0.045 | 0.680 | -0.054 | 0.576 |
| ε3/ε4*Age > 68 | 0.046 | 0.557 | -0.018 | 0.794 |
| ε4/ε4*Age > 68 | -0.445 | 0.091 | -0.389 | 0.090 |

Abbreviations: APOE = apolipoprotein E; LASI-DAD = Diagnostic Assessment of Dementia for the Longitudinal Aging Study of India; HMSE = Hindi Mental State Examination; ref = referent.

Model 1 adjusted for age (age > 68), sex (male), state of residence, top 10 genetic PCs, and *APOE* × Age > 68.

Model 2 adjusted for age (age > 68), sex (male), state of residence, top 10 genetic PCs, education (upper secondary or vocational training and tertiary education), literacy, urban/rural residence, caste, quintiles of per capita household consumption, and *APOE* × Age > 68.

Beta coefficient and p-value in bold indicates statistically significant association at p<0.05.

Asterisk (*) denotes cognitive measures for which a statistically significant interaction term was observed. Supplementary Table 11. Associations between *APOE* ε4 genotypes and cognitive measures stratified by median age.

|  | Model 1 | | | | Model 2 | | | |
| --- | --- | --- | --- | --- | --- | --- | --- | --- |
|  | **Age ≤ 68 (n = 1,361)** | | **Age > 68 (n = 1,197)** | | **Age ≤ 68 (n = 1,358)** | | **Age > 68 (n = 1,185)** | |
| **Cognitive Measure** | **Beta** | **P value** | **Beta** | **P value** | **Beta** | **P value** | **Beta** | **P value** |
| **HMSE score** |  |  |  |  |  |  |  |  |
| ε3/ε3 | ref | ref | ref | ref | ref | ref | ref | ref |
| ε2/ε3 | 0.305 | 0.478 | -0.024 | 0.964 | 0.137 | 0.716 | -0.215 | 0.676 |
| ε3/ε4 | **-0.816** | **0.009** | **-1.347** | **0.001** | -0.422 | 0.120 | **-1.242** | **0.001** |
| ε4/ε4 | -0.416 | 0.659 | -1.229 | 0.368 | -0.215 | 0.795 | -0.873 | 0.492 |
| **General cognitive function*** |  |  |  |  |  |  |  |  |
| ε3/ε3 | ref | ref | ref | ref | ref | ref | ref | ref |
| ε2/ε3 | 0.041 | 0.591 | -0.044 | 0.591 | 0.007 | 0.899 | -0.098 | 0.153 |
| ε3/ε4 | **-0.124** | **0.023** | **-0.187** | **0.002** | -0.026 | 0.501 | **-0.170** | **0.001** |
| ε4/ε4 | 0.030 | 0.857 | -0.363 | 0.082 | 0.075 | 0.519 | -0.285 | 0.095 |
| **Executive function** |  |  |  |  |  |  |  |  |
| ε3/ε3 | ref | ref | ref | ref | ref | ref | ref | ref |
| ε2/ε3 | 0.027 | 0.720 | -0.047 | 0.560 | -0.004 | 0.945 | -0.100 | 0.148 |
| ε3/ε4 | **-0.109** | **0.046** | **-0.145** | **0.013** | -0.013 | 0.748 | **-0.132** | **0.007** |
| ε4/ε4 | 0.006 | 0.969 | -0.369 | 0.072 | 0.077 | 0.542 | -0.290 | 0.091 |
| **Orientation*** |  |  |  |  |  |  |  |  |
| ε3/ε3 | ref | ref | ref | ref | ref | ref | ref | ref |
| ε2/ε3 | 0.006 | 0.927 | -0.036 | 0.637 | -0.017 | 0.747 | -0.088 | 0.211 |
| ε3/ε4 | **-0.107** | **0.022** | **-0.208** | **1.53E-04** | -0.044 | 0.248 | **-0.193** | **1.09E-04** |
| ε4/ε4 | -0.053 | 0.707 | -0.228 | 0.235 | -0.050 | 0.672 | -0.165 | 0.340 |
| **Language/fluency*** |  |  |  |  |  |  |  |  |
| ε3/ε3 | ref | ref | ref | ref | ref | ref | ref | ref |
| ε2/ε3 | 0.092 | 0.147 | 0.082 | 0.288 | 0.065 | 0.198 | 0.044 | 0.523 |
| ε3/ε4 | **-0.052** | **0.258** | **-0.153** | **0.006** | 0.011 | 0.766 | **-0.134** | **0.006** |
| ε4/ε4 | 0.120 | 0.389 | -0.185 | 0.343 | 0.124 | 0.263 | -0.149 | 0.379 |
| **Memory** |  |  |  |  |  |  |  |  |
| ε3/ε3 | ref | ref | ref | ref | ref | ref | ref | ref |
| ε2/ε3 | 0.070 | 0.395 | -0.042 | 0.638 | 0.055 | 0.447 | -0.072 | 0.391 |
| ε3/ε4 | **-0.103** | **0.085** | **-0.181** | **0.004** | -0.026 | 0.614 | **-0.175** | **0.003** |
| ε4/ε4 | -0.012 | 0.946 | -0.339 | 0.128 | 0.032 | 0.837 | -0.279 | 0.180 |
| **Visuospatial function** |  |  |  |  |  |  |  |  |
| ε3/ε3 | ref | ref | ref | ref | ref | ref | ref | ref |
| ε2/ε3 | -0.063 | 0.408 | -0.110 | 0.169 | -0.098 | 0.138 | **-0.144** | **0.046** |
| ε3/ε4 | -0.101 | 0.067 | -0.056 | 0.334 | -0.034 | 0.478 | -0.045 | 0.380 |
| ε4/ε4 | 0.151 | 0.368 | -0.257 | 0.203 | 0.148 | 0.307 | -0.200 | 0.264 |

Abbreviations: *APOE* = apolipoprotein E; HMSE = Hindi Mental State Examination; ref = referent.

Model 1 adjusted for sex, state of residence, and top 10 genetic PCs.

Model 2 adjusted for sex, state of residence, top 10 genetic PCs, education (upper secondary or vocational training and tertiary education), literacy, urban/rural residence, caste, and quintiles of per capita household consumption.

Beta coefficient and p-value in bold indicates statistically significant association at p<0.05.

Asterisk (*) denotes cognitive measures for which a statistically significant interaction term was observed.

Supplementary Table 12. Two-way interaction between *APOE* genotypes and sex on cognitive measures.

|  | **Model 1 (n = 2,558)** | | **Model 2 (n = 2,543)** | |
| --- | --- | --- | --- | --- |
|  | **Beta** | **P value** | **Beta** | **P value** |
| **HMSE score** |  |  |  |  |
| ε3/ε3*Male | ref | ref | ref | ref |
| ε2/ε3*Male | **1.446** | **0.031** | **1.420** | **0.020** |
| ε3/ε4*Male | 0.785 | 0.103 | 0.681 | 0.119 |
| ε4/ε4*Male | 2.334 | 0.133 | 1.552 | 0.269 |
| **General cognitive function** |  |  |  |  |
| ε3/ε3*Male | ref | ref | ref | ref |
| ε2/ε3*Male | 0.071 | 0.513 | 0.068 | 0.417 |
| ε3/ε4*Male | 0.106 | 0.179 | 0.082 | 0.171 |
| ε4/ε4*Male | 0.420 | 0.098 | 0.273 | 0.156 |
| **Executive function** |  |  |  |  |
| ε3/ε3*Male | ref | ref | ref | ref |
| ε2/ε3*Male | 0.065 | 0.546 | 0.074 | 0.393 |
| ε3/ε4*Male | 0.009 | 0.907 | -0.010 | 0.872 |
| ε4/ε4*Male | 0.308 | 0.222 | 0.182 | 0.362 |
| **Orientation** |  |  |  |  |
| ε3/ε3*Male | ref | ref | ref | ref |
| ε2/ε3*Male | 0.088 | 0.367 | 0.085 | 0.321 |
| ε3/ε4*Male | 0.130 | 0.064 | 0.109 | 0.075 |
| ε4/ε4*Male | 0.271 | 0.232 | 0.143 | 0.467 |
| **Language/fluency*** |  |  |  |  |
| ε3/ε3*Male | ref | ref | ref | ref |
| ε2/ε3*Male | 0.086 | 0.380 | 0.068 | 0.407 |
| ε3/ε4*Male | **0.139** | **0.048** | **0.119** | **0.043** |
| ε4/ε4*Male | 0.404 | 0.075 | 0.247 | 0.191 |
| **Memory*** |  |  |  |  |
| ε3/ε3*Male | ref | ref | ref | ref |
| ε2/ε3*Male | -0.093 | 0.432 | -0.093 | 0.390 |
| ε3/ε4*Male | 0.161 | 0.059 | **0.152** | **0.049** |
| ε4/ε4*Male | 0.371 | 0.178 | 0.286 | 0.248 |
| **Visuospatial function** |  |  |  |  |
| ε3/ε3*Male | ref | ref | ref | ref |
| ε2/ε3*Male | 0.186 | 0.089 | 0.162 | 0.093 |
| ε3/ε4*Male | 0.056 | 0.481 | 0.032 | 0.645 |
| ε4/ε4*Male | 0.322 | 0.206 | 0.248 | 0.265 |

Abbreviations: APOE = apolipoprotein E; LASI-DAD = Diagnostic Assessment of Dementia for the Longitudinal Aging Study of India; HMSE = Hindi Mental State Examination; ref = referent.

Model 1 adjusted for age, sex (male), state of residence, top 10 genetic PCs, and *APOE* × Male.

Model 2 adjusted for age, sex (male), state of residence, top 10 genetic PCs, education (upper secondary or vocational training and tertiary education), literacy, urban/rural residence, caste, quintiles of per capita household consumption, and *APOE* × Male.

Beta coefficient and p-value in bold indicates statistically significant association at p<0.05.

Asterisk (*) denotes cognitive measures for which a statistically significant interaction term was observed.

Supplementary Table 13. Associations between *APOE* ε4 genotypes and cognitive measures stratified by sex.

|  | Model 1 | | | | Model 2 | | | |
| --- | --- | --- | --- | --- | --- | --- | --- | --- |
|  | **Male (n = 1,203)** | | **Female (n = 1,355)** | | **Male (n = 1,198)** | | **Female (n = 1,345)** | |
| **Cognitive Measure** | **Beta** | **P value** | **Beta** | **P value** | **Beta** | **P value** | **Beta** | **P value** |
| **HMSE score** |  |  |  |  |  |  |  |  |
| ε3/ε3 | ref | ref | ref | ref | ref | ref | ref | ref |
| ε2/ε3 | **0.904** | **0.049** | -0.468 | 0.335 | 0.704 | 0.089 | -0.668 | 0.138 |
| ε3/ε4 | -0.498 | 0.138 | **-1.236** | **3.48E-04** | -0.370 | 0.218 | **-0.941** | **0.003** |
| ε4/ε4 | 0.470 | 0.656 | -2.093 | 0.067 | 0.324 | 0.731 | -1.317 | 0.208 |
| **General cognitive function** |  |  |  |  |  |  |  |  |
| ε3/ε3 | ref | ref | ref | ref | ref | ref | ref | ref |
| ε2/ε3 | 0.040 | 0.626 | -0.017 | 0.821 | -0.010 | 0.868 | -1.317 | 0.208 |
| ε3/ε4 | -0.079 | 0.181 | **-0.176** | **0.001** | -0.045 | 0.309 | -0.064 | 0.269 |
| ε4/ε4 | 0.075 | 0.688 | **-0.377** | **0.030** | 0.049 | 0.725 | **-0.112** | **0.006** |
| **Executive function** |  |  |  |  |  |  |  |  |
| ε3/ε3 | ref | ref | ref | ref | ref | ref | ref | ref |
| ε2/ε3 | 0.027 | 0.743 | -0.017 | 0.809 | -0.008 | 0.898 | -0.059 | 0.318 |
| ε3/ε4 | -0.103 | 0.090 | **-0.106** | **0.038** | -0.066 | 0.167 | -0.049 | 0.237 |
| ε4/ε4 | 0.006 | 0.976 | -0.310 | 0.067 | -0.005 | 0.974 | -0.181 | 0.185 |
| **Orientation** |  |  |  |  |  |  |  |  |
| ε3/ε3 | ref | ref | ref | ref | ref | ref | ref | ref |
| ε2/ε3 | 0.032 | 0.634 | -0.048 | 0.492 | -0.010 | 0.863 | -0.093 | 0.133 |
| ε3/ε4 | -0.059 | 0.231 | **-0.201** | **5.06E-05** | -0.043 | 0.311 | **-0.152** | **4.75E-04** |
| ε4/ε4 | 0.030 | 0.848 | -0.302 | 0.066 | -0.003 | 0.983 | -0.176 | 0.220 |
| **Language/fluency*** |  |  |  |  |  |  |  |  |
| ε3/ε3 | ref | ref | ref | ref | ref | ref | ref | ref |
| ε2/ε3 | 0.126 | 0.081 | 0.048 | 0.469 | 0.081 | 0.176 | 0.021 | 0.714 |
| ε3/ε4 | -0.014 | 0.796 | **-0.136** | **0.004** | 0.008 | 0.861 | **-0.088** | **0.029** |
| ε4/ε4 | 0.174 | 0.295 | -0.228 | 0.146 | 0.106 | 0.437 | -0.120 | 0.368 |
| **Memory*** |  |  |  |  |  |  |  |  |
| ε3/ε3 | ref | ref | ref | ref | ref | ref | ref | ref |
| ε2/ε3 | -0.031 | 0.719 | 0.067 | 0.417 | -0.061 | 0.430 | 0.025 | 0.736 |
| ε3/ε4 | -0.047 | 0.452 | **-0.187** | **0.001** | -0.023 | 0.681 | **-0.142** | **0.007** |
| ε4/ε4 | 0.031 | 0.874 | -0.373 | 0.054 | 0.021 | 0.905 | -0.242 | 0.166 |
| **Visuospatial function** |  |  |  |  |  |  |  |  |
| ε3/ε3 | ref | ref | ref | ref | ref | ref | ref | ref |
| ε2/ε3 | 0.023 | 0.783 | **-0.150** | **0.041** | -0.030 | 0.676 | **-0.176** | **0.007** |
| ε3/ε4 | -0.053 | 0.378 | -0.092 | 0.079 | -0.026 | 0.623 | -0.042 | 0.362 |
| ε4/ε4 | 0.139 | 0.460 | -0.215 | 0.213 | 0.150 | 0.359 | -0.143 | 0.349 |

Abbreviations: APOE = apolipoprotein E; HMSE = Hindi Mental State Examination; ref = referent.

Model 1 adjusted for age, state of residence, and top 10 genetic PCs.

Model 2 adjusted for age, state of residence, top 10 genetic PCs, education (upper secondary or vocational training and tertiary education), literacy, urban/rural residence, caste, and quintiles of per capita household consumption.

Beta coefficient and p-value in bold indicates statistically significant association at p<0.05.

Asterisk (*) denotes cognitive measures for which a statistically significant interaction term was observed.

Supplementary Table 14. Two-way interaction between *APOE* genotypes and dichotomized educational level on cognitive measures.

|  | **Model 1 (n = 2,558)** | | **Model 2 (n = 2,543)** | |
| --- | --- | --- | --- | --- |
|  | **Beta** | **P value** | **Beta** | **P value** |
| **HMSE score** |  |  |  |  |
| ε3/ε3 × > 0 years of education | ref | ref | ref | ref |
| ε2/ε3 × > 0 years of education | 0.067 | 0.913 | 0.137 | 0.821 |
| ε3/ε4 × > 0 years of education | 0.455 | 0.301 | 0.419 | 0.330 |
| ε4/ε4 × > 0 years of education | -0.070 | 0.961 | -0.467 | 0.740 |
| **General cognitive function** |  |  |  |  |
| ε3/ε3 × > 0 years of education | ref | ref | ref | ref |
| ε2/ε3 × > 0 years of education | 0.014 | 0.880 | 0.022 | 0.800 |
| ε3/ε4 × > 0 years of education | 0.052 | 0.438 | 0.055 | 0.370 |
| ε4/ε4 × > 0 years of education | -0.177 | 0.425 | -0.245 | 0.221 |
| **Executive function** |  |  |  |  |
| ε3/ε3 × > 0 years of education | ref | ref | ref | ref |
| ε2/ε3 × > 0 years of education | 0.038 | 0.692 | 0.053 | 0.554 |
| ε3/ε4 × > 0 years of education | 0.017 | 0.803 | 0.021 | 0.741 |
| ε4/ε4 × > 0 years of education | -0.123 | 0.584 | -0.189 | 0.366 |
| **Orientation** |  |  |  |  |
| ε3/ε3 × > 0 years of education | ref | ref | ref | ref |
| ε2/ε3 × > 0 years of education | -0.070 | 0.423 | -0.057 | 0.502 |
| ε3/ε4 × > 0 years of education | 3.18E-04 | 0.996 | -0.008 | 0.897 |
| ε4/ε4 × > 0 years of education | -0.035 | 0.866 | -0.086 | 0.662 |
| **Language/fluency** |  |  |  |  |
| ε3/ε3 × > 0 years of education | ref | ref | ref | ref |
| ε2/ε3 × > 0 years of education | 0.018 | 0.843 | 0.005 | 0.953 |
| ε3/ε4 × > 0 years of education | 0.059 | 0.356 | 0.052 | 0.381 |
| ε4/ε4 × > 0 years of education | -0.012 | 0.955 | -0.053 | 0.782 |
| **Memory** |  |  |  |  |
| ε3/ε3 × > 0 years of education | ref | ref | ref | ref |
| ε2/ε3 × > 0 years of education | 0.036 | 0.750 | 0.049 | 0.658 |
| ε3/ε4 × > 0 years of education | 0.087 | 0.282 | 0.103 | 0.189 |
| ε4/ε4 × > 0 years of education | -0.340 | 0.200 | -0.387 | 0.130 |
| **Visuospatial function** |  |  |  |  |
| ε3/ε3 × > 0 years of education | ref | ref | ref | ref |
| ε2/ε3 × > 0 years of education | -0.036 | 0.722 | -0.047 | 0.633 |
| ε3/ε4 × > 0 years of education | 0.071 | 0.330 | 0.077 | 0.269 |
| ε4/ε4 × > 0 years of education | -0.294 | 0.215 | -0.336 | 0.141 |

Abbreviations: APOE = apolipoprotein E; LASI-DAD = Diagnostic Assessment of Dementia for the Longitudinal Aging Study of India; HMSE = Hindi Mental State Examination; ref = referent.

Model 1 adjusted for age, sex (male), state of residence, top 10 genetic PCs, education (> 0 years of education), and *APOE* × > 0 years of education.

Model 2 adjusted for age, sex (male), state of residence, top 10 genetic PCs, education (> 0 years of education), literacy, urban/rural residence, caste, quintiles of per capita household consumption, and *APOE* × > 0 years of education.

Beta coefficient and p-value in bold indicates statistically significant association at p<0.05.

Supplementary Table 15. Associations between *APOE* genotype and cognitive measures stratified by education level (dichotomized at having received some formal schooling).

|  | Model 1 | | | | Model 2 | | | |
| --- | --- | --- | --- | --- | --- | --- | --- | --- |
|  | **0 years (n = 1,247)** | | **>0 years  (n = 1,311)** | | **0 years (n = 1,239)** | | **>0 years  (n = 1,306)** | |
| **Cognitive Measure** | **Beta** | **P value** | **Beta** | **P value** | **Beta** | **P value** | **Beta** | **P value** |
| **HMSE score** |  |  |  |  |  |  |  |  |
| ε3/ε3 | ref | ref | ref | ref | ref | ref | ref | ref |
| ε2/ε3 | 0.104 | 0.836 | 0.145 | 0.696 | -0.072 | 0.886 | 0.117 | 0.746 |
| ε3/ε4 | **-0.934** | **0.006** | **-0.685** | **0.015** | **-0.819** | **0.015** | **-0.605** | **0.026** |
| ε4/ε4 | -0.350 | 0.734 | -0.628 | 0.524 | -0.339 | 0.740 | -0.858 | 0.365 |
| **General cognitive function** |  |  |  |  |  |  |  |  |
| ε3/ε3 | ref | ref | ref | ref | ref | ref | ref | ref |
| ε2/ε3 | -0.019 | 0.746 | 0.008 | 0.915 | -0.054 | 0.355 | -0.020 | 0.747 |
| ε3/ε4 | **-0.136** | **0.001** | -0.095 | 0.081 | **-0.113** | **0.004** | -0.067 | 0.164 |
| ε4/ε4 | 0.021 | 0.865 | -0.172 | 0.362 | -0.020 | 0.867 | -0.267 | 0.108 |
| **Executive function** |  |  |  |  |  |  |  |  |
| ε3/ε3 | ref | ref | ref | ref | ref | ref | ref | ref |
| ε2/ε3 | -0.033 | 0.579 | 0.010 | 0.890 | -0.064 | 0.282 | -0.012 | 0.862 |
| ε3/ε4 | -0.094 | 0.021 | -0.083 | 0.131 | -0.074 | 0.063 | -0.056 | 0.264 |
| ε4/ε4 | -0.003 | 0.981 | -0.161 | 0.404 | -0.036 | 0.765 | -0.246 | 0.161 |
| **Orientation** |  |  |  |  |  |  |  |  |
| ε3/ε3 | ref | ref | ref | ref | ref | ref | ref | ref |
| ε2/ε3 | 0.012 | 0.857 | -0.046 | 0.425 | -0.028 | 0.672 | -0.062 | 0.265 |
| ε3/ε4 | **-0.117** | **0.010** | **-0.124** | **0.005** | **-0.092** | **0.037** | **-0.112** | **0.008** |
| ε4/ε4 | -0.052 | 0.702 | -0.079 | 0.610 | -0.079 | 0.554 | -0.125 | 0.390 |
| **Language/fluency** |  |  |  |  |  |  |  |  |
| ε3/ε3 | ref | ref | ref | ref | ref | ref | ref | ref |
| ε2/ε3 | 0.065 | 0.311 | 0.092 | 0.142 | 0.052 | 0.413 | 0.060 | 0.272 |
| ε3/ε4 | **-0.092** | **0.035** | -0.044 | 0.356 | -0.071 | 0.094 | -0.029 | 0.483 |
| ε4/ε4 | 0.047 | 0.722 | 0.039 | 0.816 | 0.016 | 0.903 | -0.050 | 0.729 |
| **Memory** |  |  |  |  |  |  |  |  |
| ε3/ε3 | ref | ref | ref | ref | ref | ref | ref | ref |
| ε2/ε3 | -0.023 | 0.752 | 0.040 | 0.637 | -0.062 | 0.401 | 0.026 | 0.750 |
| ε3/ε4 | **-0.157** | **0.002** | -0.079 | 0.218 | **-0.145** | **0.003** | -0.052 | 0.395 |
| ε4/ε4 | 0.018 | 0.904 | -0.311 | 0.164 | -0.010 | 0.949 | -0.375 | 0.080 |
| **Visuospatial function** |  |  |  |  |  |  |  |  |
| ε3/ε3 | ref | ref | ref | ref | ref | ref | ref | ref |
| ε2/ε3 | -0.072 | 0.257 | -0.105 | 0.169 | -0.083 | 0.190 | -0.136 | 0.066 |
| ε3/ε4 | -0.085 | 0.052 | -0.029 | 0.626 | -0.076 | 0.076 | -0.007 | 0.904 |
| ε4/ε4 | 0.164 | 0.213 | -0.157 | 0.443 | 0.122 | 0.347 | -0.230 | 0.236 |

Abbreviations: APOE = apolipoprotein E; LASI-DAD = Diagnostic Assessment of Dementia for the Longitudinal Aging Study of India; HMSE = Hindi Mental State Examination; ref = referent.

Model 1 adjusted for age, sex (male), state of residence, top 10 genetic PCs.

Model 2 adjusted for age, sex (male), state of residence, top 10 genetic PCs, literacy, urban/rural residence, caste, and quintiles of per capita household consumption.

Beta coefficient and p-value in bold indicates statistically significant association at p<0.05.
